# Simulation Platforms to Support Teaching and Research in Epidemiological Dynamics^⋆^

**DOI:** 10.1101/2022.02.09.22270752

**Authors:** Wayne M Getz, Richard Salter, Ludovica Luisa Vissat

## Abstract

An understanding of epidemiological dynamics, once confined to mathematical epidemiologists and applied mathematicians, can be disseminated to a non-mathematical community of health care professionals and applied biologists through simple-to-use simulation applications. We used Numerus Model Builder RAMP^®^ (Runtime Alterable Model Platform) technology, to construct deterministic and stochastic versions of compartmental SIR (Susceptible, Infectious, Recovered with immunity) models as simple-to-use, freely available, epidemic simulation application programs. In this paper, we take the reader through simulations that are used to demonstrate the following concepts: 1) disease prevalence curves of unmitigated outbreaks have a single peak and result in epidemics that ‘burn’ through the population to become extinguished when the proportion of the susceptible population drops below a critical level; 2) if immunity in recovered individuals wanes sufficiently fast then the disease persists indefinitely as an endemic state with possible dampening oscillations following the initial outbreak phase; 3) the steepness and initial peak of the prevalence curve are influenced by the basic reproductive value *R*_0_, which must exceed 1 for an epidemic to occur; 4) the probability that a single infectious individual in a closed population (i.e. no migration) gives rise to an epidemic increases with the value of *R*_0_ > 1; 5) behavior that adaptively decreases the contact rate among individuals with increasing prevalence has major effects on the prevalence curve including dramatic flattening of the prevalence curve along with the generation of multiple prevalence peaks; 6) the impacts of treatment are complicated to model because they effect multiple processes including transmission, and both recovery and mortality rates; 7) similarly, the impacts of vaccination are equally complicated and, in addition, when a fixed number of vaccination regimens are available, the rate and timing of delivery are crucially important to maximizing there ability to reducing mortality. Our presentation makes transparent the key assumptions underlying SIR epidemic models. The model and simulations tools described in this paper and the four RAMPs that we provide are meant to augment rather than replace classroom material when teaching epidemiological dynamics. Our RAMPs are sufficiently versatile to be used by students to address a range of research questions for term papers and even dissertations.

**Highlights:**

- Basic concepts used to build epidemiological models and think about epidemics are introduced
  – disease class structure and homogeneity
  – well-mixed population
  – flows of individuals among classes
  – rates of change and mathematical representation
  – deterministic versus stochastic formulations
  – disease reproductive value and R-zero
  – cessation of an epidemic versus endemicity
  – formulation of transmission
  – adaptive contact behavior
  – infectious, latent, and immunity waning periods (waiting times)
  – competing risks and rates to proportions transformations
  – effects of treatment and vaccination measures
- Principles of epidemiological dynamics are illustrated through simulation including:
  – rise to peak prevalence, subsequent fall to extirpation as herd immunity level is reached, but some individuals remain uninfected
  – level of endemicity inversely related to rate at which immunity wanes
  – effects of adaptive contact behavior on flattening the prolonging the prevalence peak
  – proportion of stuttering transmission chains that lead to an outbreak is related to size of *R*_0_ (basic reproductive rate of the disease)
  – complexities involved in incorporating treatment effects
  – trade-off between early vaccination rollout and availability of vaccination regimens
- Four simple-to-use basic and applied deterministic and stochastic runtime alterable model platforms are provided for students to use in replicating illustrative examples, carrying out suggested exercises, and exploring novel idea. These are:
  1. Deterministic SIRS RAMP
  2. Stochastic SIRS RAMP
  3. Deterministic SIRS+DTV RAMP
  4. Stochastic SIRS+DTV RAMP

## 1. Introduction

### 1.1. Motivation

The COVID-19 pandemic is a wake up call for us to be more prepared and vigilant regarding future pandemics. We can do this by enhancing our understanding of epidemic dynamics and of the most effective mitigation strategies. Part of this preparedness is providing a more sophisticated quantitative epidemiological training to students entering the healthcare and allied industries.

Many excellent text books are available for teaching epidemiological dynamics (Diekmann and Heesterbeek, 2000; Daley and Gani, 2001; Vynnycky and White, 2010; Keeling and Rohani, 2011; Li, 2018; Gudbjartsson, Helgason, Jonsson, Magnusson, Melsted, Norddahl, Saemundsdottir, Sigurdsson, Sulem, Agustsdottir et al., 2020). Some require only a beginners understanding of calculus and others a more sophisticated facility with ordinary differential equations. The most mathematically demanding require that the students have some familiarity with stochastic dynamics. This leaves the mathematically unsophisticated epidemiology student out in the cold. These students can be brought back in and provided with a heuristic understanding of epidemiological dynamics through numerical simulation. Of particular importance in this regard is hands on experience with such simulations. The roadblock here is the programming skills of students involved. The solution is to provide supportive software platforms where relative complex models can be implemented with minimal fuss and coding skills.

In this paper, we provide conceptual and simulation support for an epidemiological dynamics instruction, expecting details on specific disease to be found in other prescribed material. We begin with a review of the basic concepts and assumptions implicit in epidemic SIR (S=susceptible, I=infectious, and R=Recovered disease classes) models. We then present an epidemic simulation platform that we built using Numerus Model Builder’s RAMP™ (Runtime Alterable Model Platform) technology. Numerus RAMPs are directly downloadable from the web. Our SIRS RAMPs— we provide both deterministic and stochastic versions—though designed to help illustrate basic epidemiological concepts, may also be used to carry out simulations useful to healthcare professionals interest in exploring some aspects of pandemic management.

Our SIRS RAMPs also include treatment and vaccination classes, which allows us to discuss various aspects of treatment and vaccination strategies on epidemic dynamics. Although we discuss other extensions to SIR models that are needed to increase the utility of such models in formulating disease management policy we do not include this in the RAMPs described in this paper. We have developed other such RAMPs and web-based models for research purposes that include additional disease classes (e.g., individuals in latent, asymptomatic, symptomatic and recovery-with-some-immunity phases, or dead from the disease; (Getz, Salter, Luisa Vissat and Horvitz, 2021a) and multiple pathogen variants (Getz, Salter, Luisa Vissat, Koopman and Simon, 2021b)). Future RAMPs developed for research purposes should include demographic structure (e.g., recruitment, births, non-disease deaths, age classes), spatial structure (e.g., meta populations with some mixing, migration) and, in the context of disease ecology systems, multiple hosts with cross species transmission.

Dynamic systems models in general, and epidemic models in particular, can be deterministic or stochastic, continuous or discrete-time, compartmental systems or individual-based formulations (Getz and Lloyd-Smith, 2006; Getz, Salter, Muellerklein, Yoon and Tallam, 2018b; Railsback and Grimm, 2019). The simplest are deterministic (no random process are included) and compartmental (all individuals within the same disease class are identical and cannot be differentiated from one another in terms of when they entered this particular disease class). Most of the concept we are interested in can be demonstrated using this type of model. Deterministic formulations, however, are only suitable for modeling epidemic processes that have already taken hold in populations of tens of thousands to millions. By contrast, stochastic models are needed to computing probabilities of pathogen invasion and the probabilities that an epidemic persists when the number of infectious individuals is a tiny fraction of the population (Getz et al., 2018b). Thus besides providing a deterministic SIR RAMP as the mainstay of our presentation, we also include a stochastic compartment SIR RAMP to illustrated concepts relating to epidemic emergence and extinction. Stochastic individual-based models are needed if we want to include processes relating to length of time an individual has spent within a particular disease state (=class), or relating to other individual level phenomena. Such considerations, however, are beyond the scope of the material presented here.

### 1.2. Epidemic Modeling Concepts

Population models are idealizations of reality that include only those processes essential to addressing particular questions at hand (Getz, 1998; Getz, Marshall, Carlson, Giuggioli, Ryan, Romañach, Boettiger, Chamberlain, Larsen, D’Odorico et al., 2018a). They embody several key concepts and explicitly or implicitly include a number of assumptions that render the models a caricature of the real world. The following are a set of basic concepts and assumptions regarding the building of SIR epidemic models. The additional concepts that follow these include ways of making SIR models reflect population structures that may be critical to the utility of the model as a tool for enhancing understanding and identifying effective mitigating strategies for managing epidemics.

#### 1.2.1. Primary Concepts

##### Homogeneous population assumption

All individuals are assumed to be identical with respect to the disease process (same contact, recovery, disease-induced mortality and immunity-waning rates, as well as the same levels of susceptibility and infectivity).

##### Well-mixed population assumption

Any individual is assumed to be equally likely to make contact with any other individual in each time period (i.e., no household, healthcare, or spatial structures exist within the population that would lead to different contact rates among different groups of individuals).

##### Disease class structure

We assume that each individual belongs to one of the following three disease classes or, equivalently, be in one of the following three disease states—susceptible (S), infectious (I), and recovered with immunity (R). We may also keep track of the number of individuals that have died from the disease (D) (which can be thought of as a fourth disease state; see Fig. 1), are currently being treated (T), or have in the past been vaccinated (V). Note that sometimes we use X or Y to represent an unnamed disease class: thus in our treatment X or Y may represent, S, I, R, V, T, or D.

**Figure 1:**
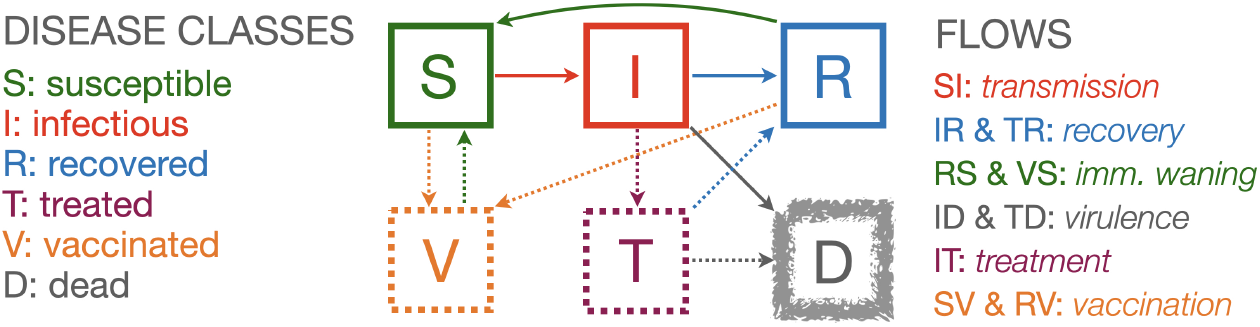
A compartmental flow diagram of an SIRS+TV epidemic process (disease classes S, I and R, the three solid squares) with the addition of deaths (D, fuzzy square) and mitigation classes (V and T, two broken squares)

##### Fundamental unit of time

We need to decide on a fundamental unit to monitor the passing of time (*t*). It is most useful if this unit corresponds to the way epidemiological data are reported, primarily new cases, hospitalizations and deaths per unit time; e.g., daily, weekly, monthly, quarterly or some other unit of time. For fast disease, such as influenza and other respiratory infections, the most useful time unit is typically a day, while for ‘slower’ diseases, such as tuberculosis or HIV, weekly, monthly or quarterly units may be more useful.

##### Variable names versus variable values

Our convention is to use roman fonts to name the disease state and italic fonts to represent the variable keeping track of the number of individuals (or density of individuals) in each disease state. Hence X is name of a state and *X*(*t*) is the number or density of individuals in this state at time, where X=S, I, R, D, T or V.

##### Flows among disease classes

Compartmental models are formulated in terms of flows of individuals from one class to the next. In particular, in many cases the per capita flows per source class are assumed constant over time, though this does not hold for the per capita flow from S to I since this will depend on the proportion of individuals that are susceptible since immune individuals are assumed to be resistant of getting infected (but see waning immunity below). We use the symbol *ρ*_XY_(*t*) to represent the per capita flow from disease class X to disease class Y at time *t* and vice versa when we are making generic statements about flow that are not disease class specific.

##### Lost immunity

Individuals in disease class R may return to disease class S because their immunity has *waned* sufficiently to render them susceptible once again to infection (see *Waning immunity* below for more details).

##### Rates of change

The change in the number of individuals in a particular disease state over a small time interval is given by the rate at which individuals enter this status minus the rate at which they leave this state over this interval of time.

##### Mathematical representation

We use *t* to denote the current time and [*t, t* + 1] to denote the up coming interval of time. In a deterministic model, using 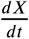 to represent the rate of change of the number of individuals in disease class X to all other disease classes, the dynamic equation for this class is given by

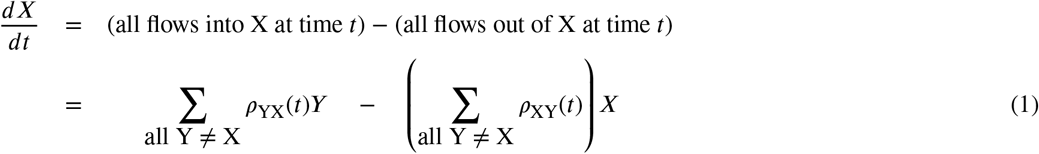

##### Deterministic versus stochastic models

If the large population is relatively large (e.g., millions) and the number of infectious individuals is at least a few hundred, then we model change deterministically in terms of the proportion of individuals that become infected over an interval of time. In smaller populations, or if we are interested in how the epidemic gets started, then we model change stochastically in terms of the probable number of individuals that become infected over an interval of time.

##### How to think about stochasticity in relation to disease class size

If with flip a fair coin many times, then as the number of times increase, so the certainty increases that half of the flips will be heads and half will be tails. If the number of flips is very large then we can ignore that fact these numbers will not be exactly 0.5 heads and 0.5 tails, because the approximation gets better as the number of flips increases. Thus when we formulate a deterministic model of an epidemic, we often remark that this model only applies for ‘large class sizes’ which means populations containing at least hundreds of thousands of individuals and hundreds of infectious individuals. In small populations, we have to account for the vagaries of probabilistic events by formulating a stochastic model. The fact that stochastic considerations are always important at the start or end of an epidemic, when the number of infectious individuals is just a few, implies that deterministic epidemic models cannot answer some critical questions about whether or not an epidemic will take off, or exactly when an epidemic may be extinguished. Stochastic models require drawing values from binomial and multinomial distributions, as will be detailed in sections 2.2 and 2.4.2.

##### Variable types

The notion that the variables *X* = *S, I, R, D, T* and *V* represent integer values is not fully compatible with the formulation of a continuous time deterministic model: in these models disease class variables actually take on continuous rather than integer values. Variables in continuous time models are, thus, more properly thought of as representing the density of individuals across undefined space of unlimited extent, which itself is a modeling idealization. In stochastic models, the values of the variables *X* are treated as integers.

##### Definition of the reproductive value/number of a disease

The reproductive value of each infectious individual in a group of similar individuals is defined to be ‘the expected number of other individuals each of these infectious individuals will go on to infect before these infectious individuals either recover or die.’ This values is highly context dependent and will change dramatically through the course of an epidemic.

##### Definition of *R*-zero (*R*_0_)

At the start of an outbreak of a new disease, when everyone except the first case (aka as ‘patient 0’ or the ‘index case’) is susceptible, the basic reproductive value is referred to as R-zero (Hethcote, 2009; Jones, 2007). The disease has the potential to become an epidemic only if *R*_0_ > 1, otherwise the outbreak will fail or just peter out after the first few cases (aka a ‘stuttering transmission chain,’ Blumberg and Lloyd-Smith 2013).

##### Definition of *R*-effective

If, initially *R*_0_ > 1, and an epidemic breaks out, then the average number of new cases that each individual infected at time *t* causes is called *R*-effective at time *t* (*R*_eff_(*t*)), which in the basic SIR model gets smaller with time, as the proportion of susceptible (S) to immune (R) individuals decreases over time. This leads to a steady decrease in *R*_eff_(*t*), from an initial value *R*_eff_(*t*) = *R*_0_ to a time where *R*_eff_(*t*) = 1. At this point, the incidence begins to decline and *R*_eff_(*t*) < 1 continues to fall because as the ratio of susceptible to immune individuals is also declining and not longer able to sustain the epidemic.

##### Cessation of an epidemic

An epidemic ceases when prevalence declines to 0, which in a deterministic model can only happen asymptotically as *t* → ∞. In stochastic models this happens in a finite time.

##### Endemicity

In populations where waning immunity causes individuals in R to become susceptible once more (i.e., transfer back to S over spending some time in R, as in the SIRS model), if this happens at a sufficiently rapid rate, then prevalence will not fall to 0 and *R*_eff_(*t*) will not necessarily decline monotonically once below 1. Rather, *R*_eff_(*t*) will either not fall below 1 or bounce back to approach 1 after falling below 1 and prevalence will stabilize at a level corresponding to a point where *R*_eff_(*t*) = 1. If this happens, the disease is said to be endemic and the approach to endemicity may exhibit dampened oscillations in prevalence levels.

##### Per capita contact rate

The SIRS model implicitly assumes that transmission can only occur through direct contact of individuals, though individuals do not necessarily need to have physical contact (e.g., short distance airborne transmission enhanced through sneezing, coughing, speaking). In the simplest SIRS models, the per capita rate *κ*(*t*) at which individuals contact others is assumed to be constant—i.e., *κ*(*t*) = *κ*_0_ for all *t*.

##### Contacts involving pathogen transmission

Transmission only occurs when a susceptible and infectious individual contact one another. The per capita rate at which any individual in the population contacts an individual in the infectious class, is the contact rate *κ*(*t*) multiplied by the proportion *I*/*N* of infectious individuals in the population—i.e., *κ*(*t*)*I*/*N*. Thus the total rate of contact is the number of susceptible individuals *S* multiplied by the per capita rate, which thus is *κ*(*t*)(*SI*/*N*). Only a proportion of these contacts will lead to actual transmission, as discussed next.

##### Risk of transmission

Transmission is best thought of as the probability of a sufficiently high dose of pathogens being transferred from infectious individual to a susceptible individual during an event that is characterized as a contact to cause disease in the susceptible individual. Although transmission is highly complicated, depending on how close individuals are and for how long they make contact during a ‘contact event,’ these complexities are washed out through an averaging processes encapsulated in a ‘risk or force of infection’ parameter most often referred to as beta (represented by the Greek letter *β*). In an SIRS model the actual per capita rate of transmission is thus a concatenation of processes involving contact rates and probabilities of transmission per contact with an infectious individual (this computation is characterized more precisely in section 2.2). Thus the transmission rate itself is represented by the product *βκ*(*t*)*I*(*t*)/*N*(*t*). In most SIRS models, contact rates are often folded into the value of *β*, thereby leading the disappearance of the parameter *κ*.

##### Frequency dependent transmission

The per capita contact rate is a constant (i.e., independent of population size) *κ*(*t*) = *κ*_0_, implying the per capita contact rate of any susceptible with an infectious individual is *κ*_0_*I*(*t*)/*N*(*t*), so the total transmission rate at time *t* is

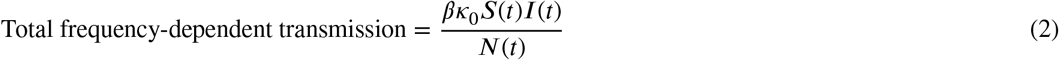

##### Mass action transmission

If the per capita contact rate scales with population density *N*, as would be the case if individuals moved around at random making contact as they bump into one another in what is referred to as a ‘mass action process’, then we need to replace *κ*_0_ in Eq. 2 by *κ*_0_*N* to obtain

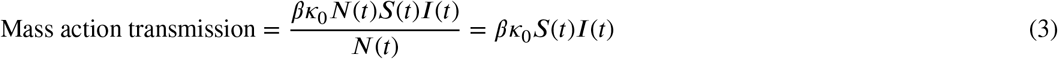

This type of transmission process only applies to entities that have no autonomy over their movement as the move and bounce around when contacting one another.

##### Adaptive (behavior modified) contact

In more sophisticated SIRS models, a behavioral response can be included by assuming that *κ*(*t*) depends, say, on the current prevalence *I*(*t*)/*N*(*t*). We refer to this as an adaptive contact rate because the rate adapts to the level of disease prevalence in the population so as to reduce transmission. In this case we replace the constant basic rate, with an expression that decreases in value with increasing levels of the prevalence *I*/*N* (see section 2.1.4 for a formulation of one such function).

##### Waiting times and relationship to transition rates

In SIRS models, if *ρ*_XY_ is the rate at which individuals in disease class X flow (aka transition) to disease class Y, then it can be shown (Box 1) that the expected time spent in disease class X (aka waiting or residence time in disease class X) when the only path out of X is to disease class Y, is given in our notation (See Box 2) by 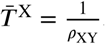. Naturally, waiting times are reduced when more then one process exists for exiting disease class X, as discu^*ρ*^s^X^s^Y^ed in Box 2.

##### Infectious period

The infectious period is the waiting time in infectious class I.

##### Latent period

In some disease models a latent period is included in which it is assumed that an individual has been exposed to a pathogen and enters a class denoted by E = exposed or, in our formulations by L = latent, before entering infectious class I once the latent period is over. (Such extended models are often referred to as SEIR models). The latent period is the waiting time in latent class L (or E in SEIR models). The addition of a latent period does nothing to alter the epidemic dynamics (e.g., does not influence *R*_eff_) other than to introduce a time-delay into the rate at which the epidemic breaks out.

##### Disease-induced mortality (virulence)

In some SIRS models, individuals in infectious class I can either recover or die (i.e., the flow out of I is into both R and D, as indicated in Fig. 1)

Students may be introduced to some of these additional concepts, as appropriate to the material presented in instructional settings, although these concepts are not implemented in the deterministic (continuous time) or stochastic (discrete time) SIRS+DTV RAMPs considered here. Other current (Getz et al., 2021b) and possible future SIRS-RAMPs will include some of these concepts.

#### 1.2.2. Additional Concepts

##### Disease classes beyond SIR

Additional intrinsic disease classes (i.e., other than those associated with interventions) include, as already mentioned, infected but not yet infectious (L = latent, aka E = exposed), as well as infectious without symptoms (A = asymptomatic), or contacted (C) but not necessarily on the way to becoming infectious (i.e., individuals in C may either go back to S or onto L) (Getz et al., 2021a). Class A is extremely important in masking the seriousness of an outbreak, as we have seen with the recent COVID-19 pandemic (Park, Cornforth, Dushoff and Weitz, 2020; Gao, Xu, Sun, Wang, Guo, Qiu and Ma, 2021). Class C allows for considerations of the contact tracing and subsequent quarantining of individuals who may or may not then go onto to become infectious (Ferretti, Wymant, Kendall, Zhao, Nurtay, Abeler-Dörner, Parker, Bonsall and Fraser, 2020; Yasaka, Lehrich and Sahyouni, 2020).

##### Waning immunity

Individuals are assumed to be immune for a time after recovering from infection (i.e., once no longer infectious). This immunity wanes over time with individuals becoming increasingly susceptible over time. In SIRS models, however, the implicit assumption is that individuals in disease class R transfer back to disease class S at a constant rate, irrespective of how long any particular individual has spend in disease class R. In SIRS compartment models, the average time spent by any individual in disease class R, however, is the so-called disease class R waiting time. In reality, it may take a higher dose of pathogen to infect individuals a second time.

##### Breakthrough infections

These are defined to be infections in individuals that have been vaccinated (Schieffelin, Norton, Kolls et al., 2021), but this notion could equally well apply to individuals that have been previously infected. In compartmental models, flows from R and V back to S allow us to account for breakthrough infections at a very crude level because they do not take the subtleties of waning immunity into account for individuals in R or V. A refined treatment requires an individual-based or agent-based approach as described below (also see Getz et al. 2021b for including of breakthrough infections in a multi-variant pathogen setting).

##### Demographic class structure

Transmission and mortality rates may often be age dependent. For example, the global influenza pandemic of 1917-1919 proved to be most virulent for young adults while the COVID-19 pandemic was most lethal for the elderly. When age-structure is added to a model then a ‘mixing matrix’ is needed to describe the relative frequencies with which individuals of different ages make contact with one another (Glasser, Feng, Moylan, Del Valle and Castillo-Chavez, 2012). Other demographic classes may be job related, such as partition off healthcare workers (Lloyd-Smith, Galvani and Getz, 2003) or teachers for special consideration.

##### Metapopulation structure

The spatial homogeneity assumption implicit in SIRS models can be obviated by dividing a population into a number of sub-populations, each of which is itself homogeneous, but a mixing/movement matrix is needed to describe how individuals in one sub-population contact or join individuals in another sub-population (Keeling, 1999; Lloyd and Jansen, 2004; Watts, Muhamad, Medina and Dodds, 2005; Ajelli, Gonçalves, Balcan, Colizza, Hu, Ramasco, Merler and Vespignani, 2010).

##### Epidemic versus demographic time scales

The basic SIRS model does not include rates at which new individuals join a population (aka recruitment), are born into a population or die from natural causes. If an epidemic occurs on a time table that is fast compared with the demographic processes of births, maturation of individuals (as in recruitment to a sexually active population in the context of sexually transmitted diseases) or deaths, then these demographic processes can be ignored (Hastings, 2010). In the case of ‘fast disease’ such as influenza in humans, the epidemic may often burn through the population in months, while births and deaths only lead to significant changes population numbers on the scale of decades. For slower diseases such HIV-AIDS, tuberculosis, or leprosy, the epidemic process may be in the same time scale as the demographic time scales as births an natural deaths, particular in species that have generation times measure in years rather than decades. Beyond epidemic and demographic time scales, longer evolutionary time scales may also be considered (Feng, Smith, McKenzie and Levin, 2004).

##### Multiple hosts

In zoonotic diseases transmitted from animal to human hosts, such as certain strains of influenza and cold viruses, it may be important to consider how the pathogen is transmitted back and forth between human and animal populations (Dobson, 2004; McCormack and Allen, 2007). This is certainly the case for avian influenza (Gumel, 2009).

##### Multiple pathogen variants

During the course of an epidemic the pathogen may evolve, as occurred in the recent COVID-19 pandemic. In this case, particularly if some variants are much more transmissible or lethal than others, models that incorporate multiple pathogen strains may be used to help manage strain proliferation (Getz et al., 2021b).

##### Other modes of transmission

SIRS models are applicable to directly transmitted diseases and are generally not applicable to pathogens that are transmitted by a vector (e.g. tick or mosquito borne diseases) when the pathogen in the vector population varies over time. In this case, the pathogen levels in both populations need to be modeled, as well as the rates of contacts between hosts and vectors and how these contacts may vary over time (Dobson, 2004; Cai and Li, 2010). Beyond vector transmission there are various environmental modes of transmission including water borne (Tien and Earn, 2010) and soil borne diseases (Gilligan, 1990).

##### Agent-based models

ABMs, also known in some contexts as IBMs (individual-based models) allow us to focus on the history of individuals (Getz, Gonzalez, Salter, Bangura, Carlson, Coomber, Dougherty, Kargbo, Wolfe and Wauquier, 2015a; Railsback and Grimm, 2019). Among other things, this means we can directly account for the time that each individual entered a particular disease state and make the exit from that state dependent on how long the individual has been in that state. This individual level control obviates the exponential distribution of waiting times in diseases implicit in compartmental systems models (Boxes 1), although a somewhat cumbersome ‘boxcar’ extension to compartmental system models allows us to obtain waiting-time distributions that have a humped-shaped distribution with the mode close to the mean (see Appendix B). Such hump-shaped distributions also arise in ABMs if the rate individuals leave a particular disease class increase with the time they have spent that class (see Appendix C).

## 2. Methods

### 2.1. SIRS Epidemic Model Formulation

#### 2.1.1. Basic model

The continuous time, extended Kermack and McKendrik SIRS (S=susceptibles, I=infected & infectious, R=recovered) epidemic model (Kermack and McKendrick, 1927; Li, 2018) is foundational to the development of epidemiological systems modeling and theory (Hethcote, 2000). It is most often formulated in terms of the variables *S, I*, and *R* (note we use the roman font mnemonics S, I, and R to refer to the names of the disease classes/compartments and italic fonts to the variables themselves). For the sake of completeness we include a disease class D to account for the dead, but define the number of individuals alive at time *t* to be

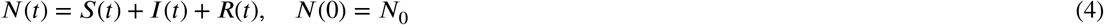

In the context of differential equation models these variables, strictly speaking, should be interpreted as population density variables; though loosely speaking we think of them in terms of numbers in a closed (no births or natural deaths, no migration), homogeneous population (everyone is equally susceptible, infectious, and likely to make contact with any other individual). From the general systems description given by Eq. 1, if the flow of individuals from disease classes/compartments S to I, I to R and R back to S, plus a flow of individuals from I to D (disease-induced mortality process) are respectively represented by the per capita flow rates *ρ*_SI_, *ρ*_IR_, *ρ*_RS_, and *ρ*_ID_, (solid arrows in Fig. 1), then we obtain the deterministic continuous-time epidemic compartmental SIRS model

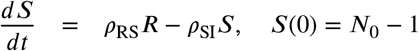

##### Box 1

**SIRS exponential transfers mean period**

Focusing purely on the flow of a cohort of individuals in disease class X at time *t* = 0, which we denote by *X*_0_, if the constant per capita outflow rate is *ρ*, then the number of individuals in this cohort that still remain in *X* at time *t* is given by the differential equation

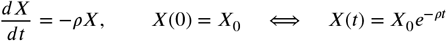

Note that we loosely think in terms of numbers of individuals because *X*(*t*) takes on continuous rather than integer values. Strictly speaking, it is more accurate to think of the proportion of individuals that still remain in disease class X at time *t* as being given by 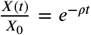, because proportions are not required to be integer values and, in the case of a single individual, this value represents the probability that an individual in disease class X at time 0 is still in disease class X at time *t*. This also implies that the probability that an individual in disease class X leaves this class by time *t* is given by the cumulative probability function

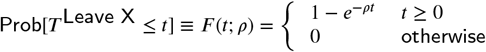

The corresponding probability density function is

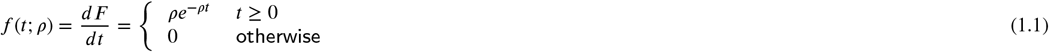

By computing 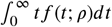 we obtain the mean time each individual is expected to spend (i.e., waiting or residence time) in class X as

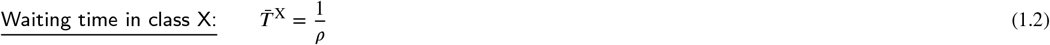

##### SIRS+D cont.-time deterministic model

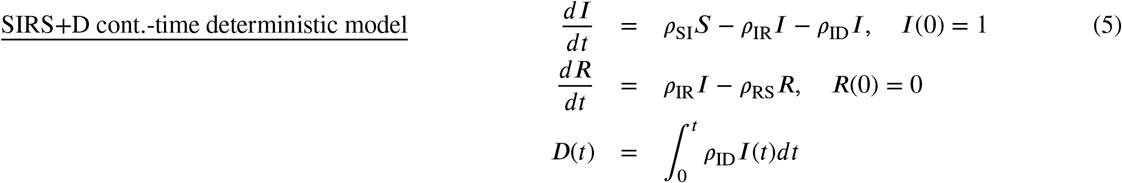

We note the initial conditions included here reflect the start of an epidemic caused by a novel pathogen (and variables, though actually continuous, are interpreted in terms of numbers of individuals). Other initial conditions can be assumed when using Eq. 5 to simulate a population that is already part way into an epidemic.

In practice, once the values or functional forms of the flows *ρ*_XY_ have been specified for X, Y = S, I and R, solutions will be generated over the time interval [0, *t*_fnl_] using numerical simulation techniques.

#### 2.1.2. Constant flow rates

In the simplest case, apart from the per capita susceptible flow rate *ρ*_SI_, the flows are assumed to be constant and the transfer of individuals from one disease state to another follows an exponential process. The consequences of this assumption are discussed in Box 1.

##### Extra information

The assumption of a constant flow through X, resulting in an exponential exit rate for all members currently in X, is unrealistic. To see this, consider any particular individual entering X at time *t* = 0. The probability this individual leaves X during the interval [*t, t* + 1] is given by (recall Eq. 1.1 in Box 1)

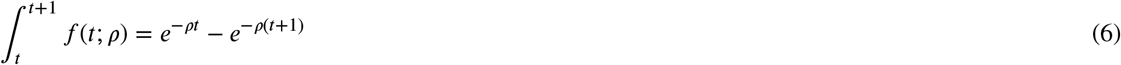

This probability has a maximum value of 1 − *e*^*ρ*^ at *t* = 0. If individuals spend several units of time in X, it is much more reasonable to assume that the probability they leave X in any particular unit of time is around the mean time 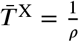 spent in X rather than the very first period of time. One can achieve this more reasonable departure behavior in a continuous-time differential equation model by dividing the disease class X into a say *K* > 1 sub-compartments and have an individual pass through each of these sub-compartments at a rate *Kρ*, resulting in so-called box car models (Appendix B). The exit process from a box-car model with *K* compartments is by an Erlang distribution with shape parameter *K* and scale parameter *Kρ* rather than and Exponential distribution with scale parameter *ρ*. In an Erlang distribution for which *K* > 1, an individual is most likely to leave during the period that surrounds the mean time that it spends in X, which is 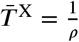, rather than immediately on entering, as implied by Eq. 6 for the exponential case. The box car or Erlang model is des^*ρ*^cribed in more detail in Appendix B.

#### 2.1.3. Disease transmission

The per capita flow rate *ρ*_SI_ from the susceptible to the infectious class is more complicated than the per capita flow rates between the other disease classes. This flow rate represents a disease transmission process that depends on susceptible and infectious individuals making contact with one another. This contact process in the original Kermack-McKendrick model was assumed to be dependent on the product *S*(*t*)*I*(*t*) of the susceptible and infectious classes. This approach draws its inspiration from the *Law of Mass Action* that pertains to chemical kinetics. The resulting incidence— the rate of new infections—then takes on the eponymous *βSI* transmission rate, where *β* is a parameter whose value reflects the joint effects of both an underlying contact rate and a ‘probability of transmission’ per contact (inverted commas are used because only in the stochastic model, as developed in section 2.2, are probabilities made explicit; also see section 2.1.5 on how to derive the probability of transmission per contact). A more appropriate approach to modeling disease transmissions, and hence the incidence rate, in humans (at all but the very lowest population densities found in rural areas) is to assume that: 1.) each individual in the population contacts others at a per capita rate *κ*; and 2.) only the proportion of contacts with infectious individuals—i.e., *I*(*t*)/*N*(*t*)—can lead to transmission. Further, not every contact will lead to transmission, but will be scaled according to a *force of infection* parameter *β*. In this case, we typically represent transmission by a per capita susceptible function that has the *frequency dependent* form (Getz and Pickering, 1983)

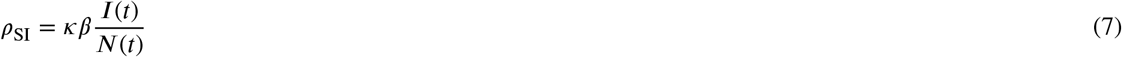

In this case, total incidence, which we denote by Δ^+^*I*(*t*) (Δ^+^, is used to denote change due to new additions to I, before accounting for the individuals that leave I), is now given the function

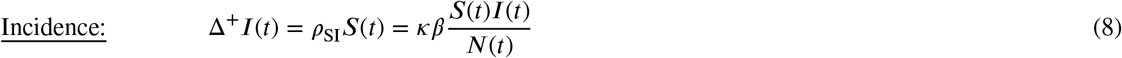

More elaborate incidence functions have been proposed (McCallum, Barlow and Hone, 2001), particularly in the disease ecology literature (Dougherty, Seidel, Carlson, Spiegel and Getz, 2018). Eq. 7, however, is the one we will focus on throughout this exposition. Also, in most of the literature, the processes of contact, represented by the parameter *κ* and force of transmission per contact, represented by the parameter *β*, are not separated but are effectively rolled into a single constant *β*_*κ*_ = *κ* × *β*, with no subscript used (equivalent to setting *κ* = 1). In this case, the contact process itself is no longer transparent. We believe that the contact process should always be explicit as a reminder that transmission requires ‘effective contacts’ to be made. Note that it does not make SIR models dynamically more complex when *β*_*κ*_ is represented as the product of two constants: i.e., when *β*_*κ*_ = *κ* × *β*. An effective contact is one that result in pathogen transmission. Contacts that do not result in transmission are part of more complex epidemic models that consider quarantining as a mitigation strategy (Getz et al., 2021a) because not all quarantined individuals become infected.

#### 2.1.4. Constant and adaptive contact rates

Most SIR models implicitly assume a constant contact rate

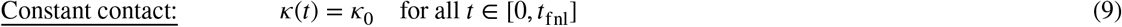

Arguably, one of the most important modifications to the transmission rate and incidence expressions given by Eqs.7 and 8 is to account for changes in contact rates once a pandemic has started. This behavior was seen with regards to the Ebola pandemic of 2015 (Getz et al., 2015a), as well as COVID-19 pandemic (Getz et al., 2021a). One way to deal with this is to assume the contact rate adapts, by decreasing either as a function of incidence (Eq. 8) or the proportion of individuals that are infectious (i.e., *I*(*t*)/*N*(*t*)). If we assume the latter, then a three parameter form for the contact rate *κ*(*t*), that depends on a basic contact rate *κ*_0_ > 0, an infectious proportion switching point *P*_*κ*_ ∈ (0, 1), and an abruptness switching parameter *σ*_*κ*_ *≥* 0 is given by the function (Getz et al., 2021b):

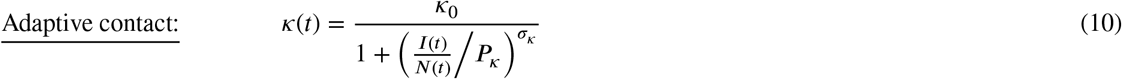

The switching point parameter *σ*_*κ*_ may be set to 2 for gradual switching, 5 for relatively abrupt switching, or 20 to approximate a step function around the infectious proportion switching point *P*_*κ*_.

#### 2.1.5. The basic reproductive rate R-zero and probability of infection

The basic reproductive rate, *R*_0_, of a disease at the start of an epidemic in a population that has never been exposed to the pathogen causing the outbreak represents the number of expected new cases that the index case (aka patient zero) will cause. This number is represented by

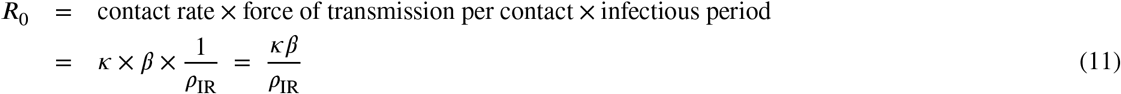

The product *κβ* can be used to obtain the incidence per susceptible over a short time interval Δ*t* at the start of the epidemic (also interpreted as the probability of a susceptible becoming infected over [0, 1]) by solving for the incidence over the period *t* ∈ [0, Δ*t*], assuming *I*_0_ = 1. This number can be obtained by first solving the differential equation

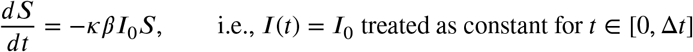

Integrating the above equation for the condition *I*_0_ = 1 (to see how many susceptibles are infected by a single inectious individual; and assuming both *κ* and *β* constant over this interval) implies that *S*(Δ*t*) = *e*^−*κβ*Δ*t*^*S*_0_. Since the expected proportion of susceptibles that will become infected over [0, Δ*t*] (i.e., those that leave class S for class I) is given by 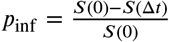 (also interpreted as the initial probability of infection), we obtain the expression

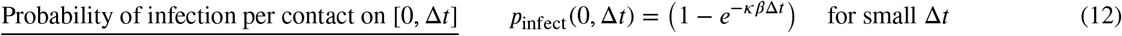

Finally, we reiterate that most SIRS formulations do not explicitly identify a contact rate *κ* so that this parameter does not appear at all in these formulations and also that *γ* is used instead of *ρ*_IR_ (*γ* ≡ *ρ*_IR_). In such cases, *R*_0_ = *β*/*γ* and *P*_*infect*_(0 Δ*t*) = (1 − *e*^−*β*Δ*t*^). But, as previously mentioned, it is useful to explicitly include *κ* because it is needed to discuss adaptive contact and provide the structure needed to introduce quarantine rates in a sensible way.

### 2.2. SIRS Discrete and Stochastic Formulations

The discretized version of the SIRS model when disease-induced deaths are not considered is given by the following set of *t* = 0, 1, …, *t*_fnl_ when simulated over the interval [0, *t*_fnl_]

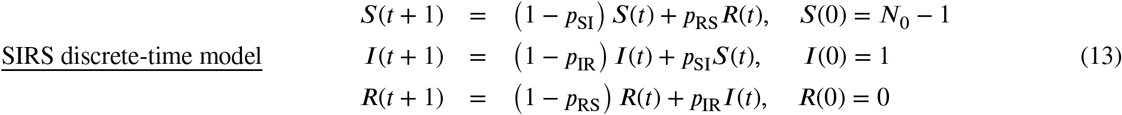

where the proportions *p*_*XY*_ are related to the rates *ρ*_*XY*_ by the equation (see Box 1 and cf. Eq. 12)

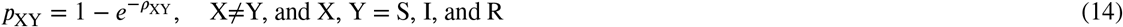

Thus, as required, *p*_XY_ = 0 when *ρ*_XY_ = 0 and *p*_XY_ → 1 as *ρ*_XY_ → ∞.

A discrete-time stochastic version of this model is obtained from Eq. 13 by assuming that the quantities *p*_SI_ represent probabilities proportions. (Note: Continuous-time stochastic models require a level of mathematical treatment beyond the scope of this presentation; e.g, see Allen 2017). In this case, the simulation takes the form of a discrete-time Monte Carlo process in which drawings from a binomial distribution are considered rather than computations of proportional change. Specifically, simulation involves the following drawings, using the following notation to denote such drawings (aka samplings):

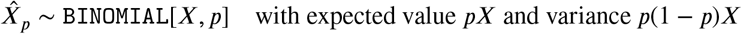

We note that 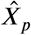 is the actual number of actual objects drawn or sampled from a total of *X* objects, where each object has the probability *p* of being selected (and hence 1 − *p* of not being selected).

Formally, our stochastic model is the following Monte Carlo simulation that begins with specified values for *S*(0), *I*(0), *R*(0) and *D*(0) and continues with successive samplings for *t* = 1, 2, …, *t*_fnl_ :

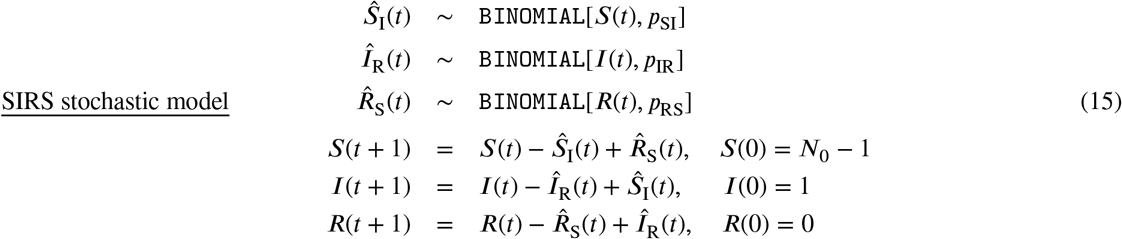

### 2.3. SIRS+D Stochastic Formulation

The discrete-time and stochastic models given by Eqs. 13 and 15 do not include disease-induced mortality (aka mathematical epidemiologists as virulence). Thus they are only useful for considering the start or initial growth of epidemics or epidemics where disease-induced mortality is inconsequential. Including deaths requires consideration of competing rates/risk for individuals leaving particular disease classes. For example, when deaths are considered individuals leave class I to either recover (R) or die (D). In this case, a competing rates formulation (Box 2) is needed to simulate outcomes. In particular, in the SIRS+D continuous-time model (Eq. 5) the prevalence equation (*I*(*t*)) involves the competing rates of recovery (*ρ*_IR_) and disease-induced mortality (*ρ*_ID_). Thus applying a competing rates formula, we obtain (Box 2, Eq. 2.3 with Δ*t* = 1)

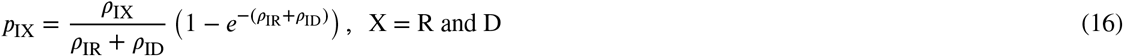

Since individuals in the prevalence class I are now faced with two possibilities of where to go—i.e., recover or die—the stochastic version of the model requires that the binomial drawing be replaced with a multinomomial drawing that depends on the two probabilities *p*_IR_ and *p*_ID_. This requires that we use a multinomial function that, in general, for the case of *n* − 1 leaving options with competing probabilities *p*_*i*_, *i* = 1, …, *n* − 1, and not to move at all with probability 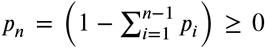. We use the following convention to specify the values obtained in one such multinomial sampling:

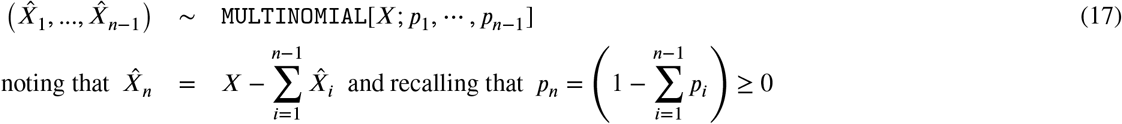

Thus the model is now given by (cf. Eq 15)

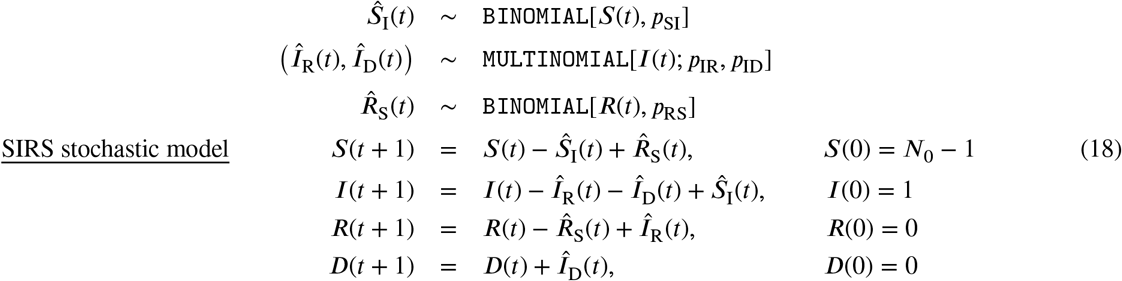

### 2.4. SIRS+DTV: Including Treatment and Vaccination

We now add treatment (T), and vaccination (V) classes to our SIRS+D model and define

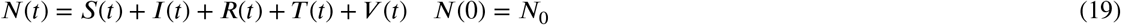

#### Box 2

**Competing rates/risk formulation**

Consider a compartmental system with *J* different classes and assume individuals leave class *X*_*i*_ to enter class *X*_*j*_ at a per capita rate *ρ*_*ij*_, *j* ≠ *i, i* = 1, …, *J* (e.g. leaving an infectious class due to recovery or death), then we have the system of equations

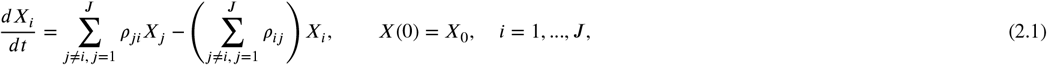

##### Proposition

Under the assumption that the rates *ρ*_*ij*_ are constant over the interval [*t, t* + 1] and using the following:

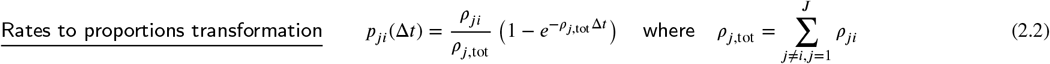

the solution to Eq. 2.1 over the time interval [*t, t* + 1] is given by the ‘competing rates’ expression

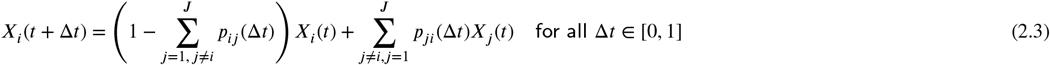

##### Verification

It follows from expression Eq. 2.2 that

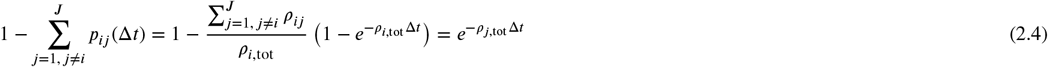

Substituting expressions Eq. 2.2 into Eq. 2.3, rearranging terms and dividing by Δ*t* we obtain

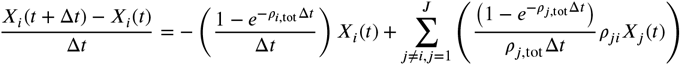

Expanding the exponential functions leads to

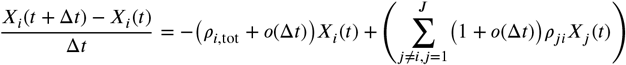

Therefore, in the limit as Δ*t* → 0, we obtain Eq. 2.1 when using the second expression in Eq. 2.4.

##### Waiting times

Since the flow rate of individuals out of disease class X is now given by 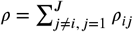, from Eq. 1.2 in Box 1 it follows that the waiting time of individuals in class X_*i*_ is given by

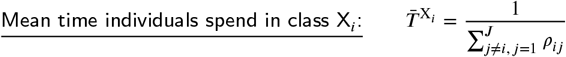

For simplicity, we assume that individuals under treatment are no longer able to transmit pathogens to the population at large—i.e., they are essentially completely quarantined. In reality, however, the situation is much more complicated than this: the ability of individuals under treatment to transmit pathogens depends on how strict the quarantine procedures are. In addition, if we structure the population to include healthcare workers or members of the infected individuals household, then clearly these healthcare workers and household members are likely to be at some particular risk of infection from individuals under treatment. Thus, moving beyond our simplifying assumption of individuals under treatment being effectively completely quarantined requires additional model structure and transmission parameters, which is not included in the model formulated here.

#### 2.4.1. Deterministic Formulation

The dynamic equations consist of the following system of 5 differential equations, augmented by the three integrations that allow us to keep track of the accumulating deaths (*D*(*t*)), cases that are treated (*T* ^total^(*t*)), and individuals that are fully vaccinated (*V* ^total^(*t*))

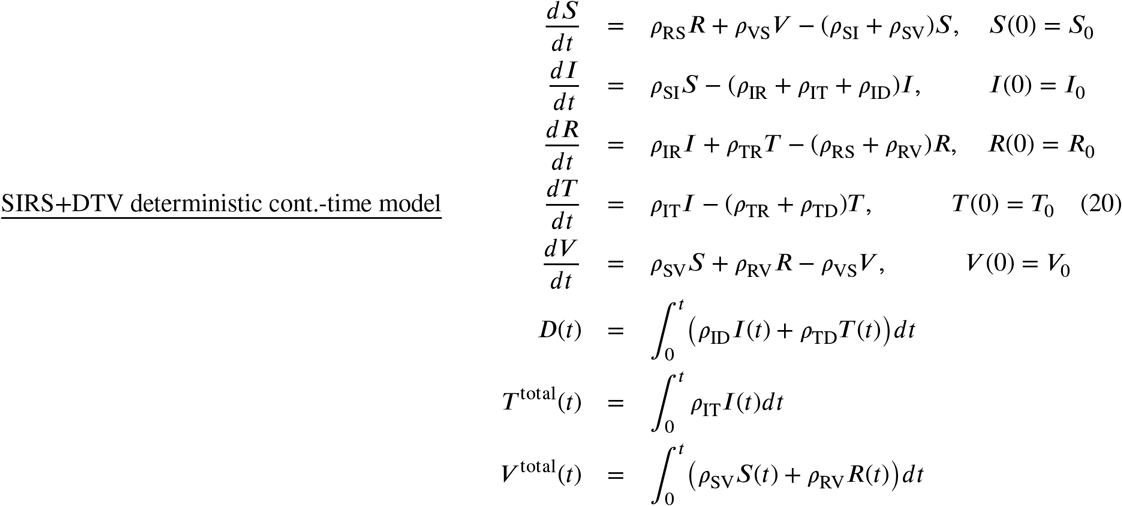

For simplicity, we will initially assume a constant treatment rate *ρ*_IT_ = *ρ*_treat_ that is implemented from the onset of the epidemic being modeled. However, we will place a maximum *T*^max^ on the number of individuals that can be in treatment at anyone time, due to a limited capacity of the healthcare system to take care of sick individuals. Thus, we have

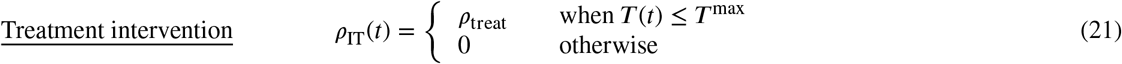

This intervention, however, will be available to the user to elaborate through a RAM (runtime alternative module) that is the hallmark of our RAMP (runtime alterable model platform) technology. Eq. 21 will be the `Default` formulation while alternative 1 will be constant application of *ρ*_IT_ with not upper limit.

For the sake of completeness, we note that since the outflow from I now includes both flows to R, T, and D the expression for *R*_0_ in Eq. 11, while individuals in T and D are assumed not to transmit pathogen (note for disease, such as Ebola, pathogen is transmitted from individuals in D preparation of the corpses for burial; Akwa and Maingi 2020) now becomes

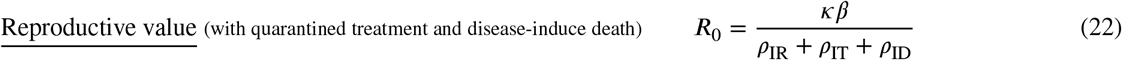

For simplicity, we will initially assume a constant vaccination rate *v* that is implemented from time 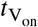 onwards. As with treatment, we will place an upper bound on the number of vaccination regimens that can be administered, where a vaccination regimen is defined to be a complete course that consists of one or more shots over a specified interval of time. Note, for simplicity, we assume that the full effect of the vaccination starts at the time of administering the first shot in the prescribed regimen (at which time the individual is transferred to V from S, C, L, R, or Q, as the case may be). An individual that later transfers from R back to S may then receive a second regime in an ongoing vaccination rollout program. The vaccination rate *v* itself will be available to the user to elaborate through our Vacc RAM (runtime alternative module). Thus our default vaccination rollout program is defined by

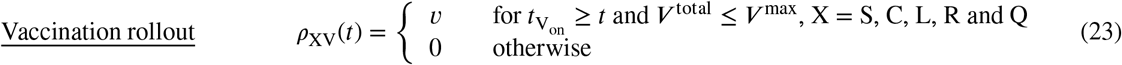

This function will be alternative 1 of the `Vacc` RAM of our RAMP.

#### 2.4.2. Stochastic Formulation

For each time step in the stochastic simulation (*t* = 0, 1, 2, …, *t*_fnl_ −1), we need to transform all the rates *ρ*_XY_ (*t*) (used in the continuous-time formulation) to probabilities *p*_XY_(*t*) using the *rates to proportions transformation* Eq. 2.2 in Box 2. We then proceed at each time step to use these probabilities in the Monte Carlo drawings followed by computing next values using the updating equations, as specified below:

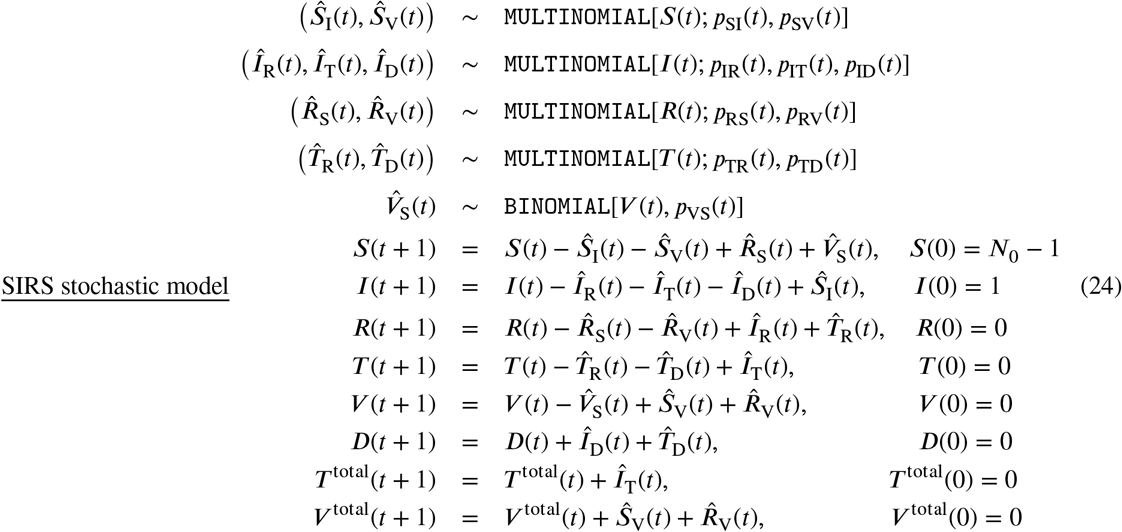

Once the simulation is complete, we will have generated time series for all the disease class variables *X*(*t*), *t* = 0, …, *t*_fnl_, *X* = *S, I, R, T* and *V*, as well as an incidence time-series *Ŝ*_*I*_ (*t*) and new deaths time-series 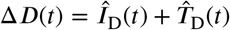 over *t* = 1, …, *t*_fnl_.

### 2.5. RAMP Implementation

Here we provide a brief description of the structure and operation of our four RAMPs, each of which is an HTML file the implements the reference models as a Numerus Model Builder RAMP application

**SIRS_Det.xml** implements the continuous-time deterministic model represented by Eq. 5, with *ρ*_ID_ fixed at 0

**SIRS_Sto.xml** implements the discrete-time stochastic model represented by Eq. 15

**SIRS+DTV_Det.xml** implements the continuous-time deterministic model represented by Eq. 20

**SIRS+DTV_Sto.xml** implements the discrete-time stochastic model represented by Eq. 24

SIRS+DTV continuous-time deterministic and discrete-time stochastic RAMPs, while the structure and operations of our simpler SIRS continuous-time deterministic and discrete-time stochastic RAMPs are just a subset of descriptions of the more complicated RAMPs once elements relating to death (D), treatment (T) and vaccination (V) processes are removed. The RAMPs are built with the Numerus Model Builder Designer (**NMB Designer**). Once built the **NMB Designer** is used to generate a RAMP as an HTML file with a user provided name. This file can then be read and implemented by using **NMB Studio**. This application is available for free at the Numerus Website (URL). The reader and the RAMP HTML files for the continuous-time deterministic and discrete-time stochastic SIRS+DTV RAMPs are available at our Numerus Model Builder Website, where more information on using these RAMPs is available at the Numerus Website.

#### 2.5.1. Deterministic RAMP dashboard

Once **NMB Studio** has been download from the Numerus Model Builder Website, installed, launched, and the Continuous-time deterministic SIRS+DTV RAMP HTML file has been read in, the dashboard depicted in Fig. 2 appears. It is used to load saved or reset parameter values for simulations, initiate runs, generate graphs, and save simulation results in the form of CSV files. We note that every object on the dashboard (parameter sliders, RAM rollers, and graphs) has a string of three alphanumeric characters (which we refer to as airport codes) associated with it that is used in scripts to control parameter values, select runtime alternative module (RAM) options, and select among graphing options. The various dashboard windows and their functions are:

**Figure 2:**
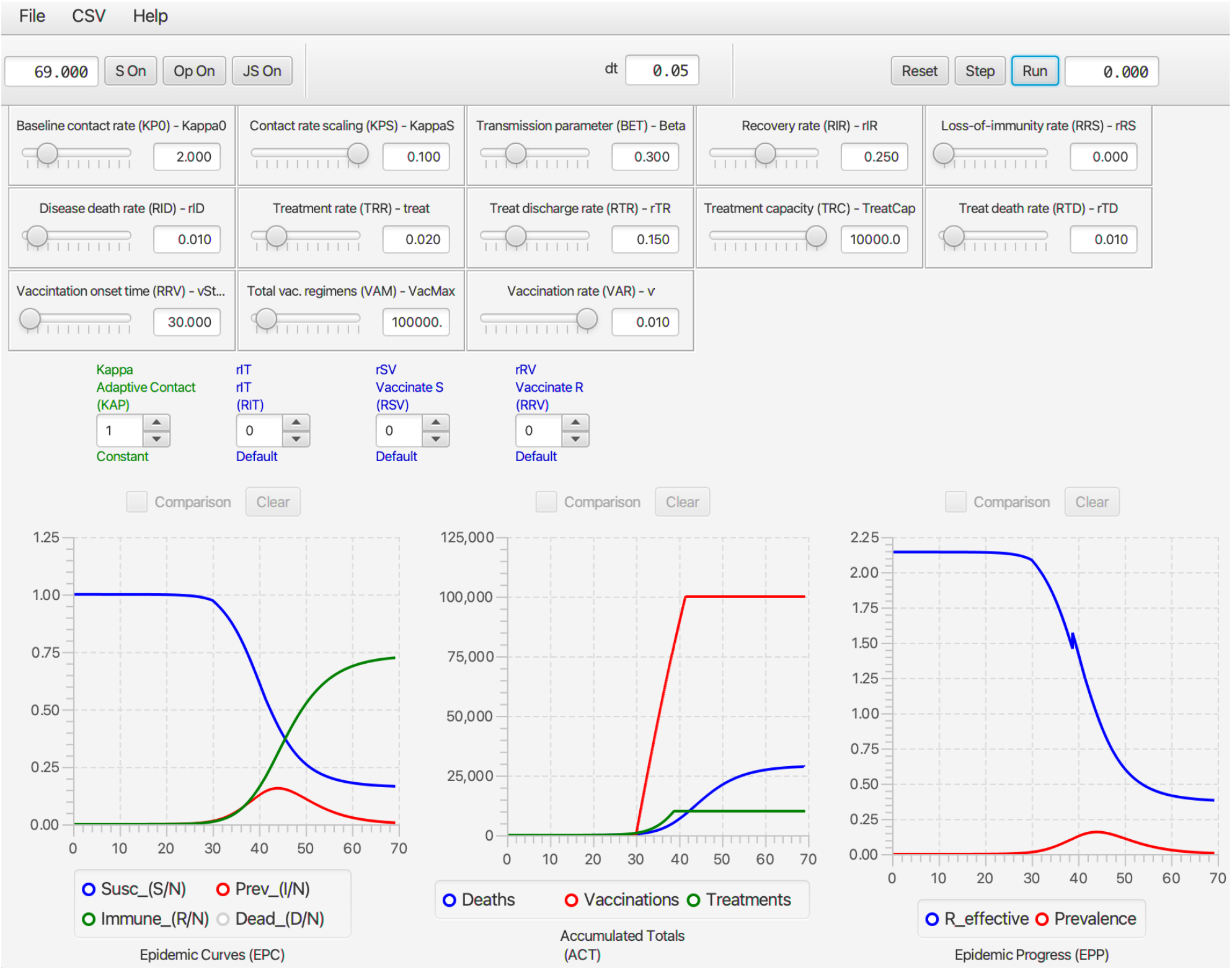
The dashboard of the SIRS+DTV continuous time RAMP is depicted here with graphical results of a run made over the interval [0,70] using the set of values appearing in the windows of the 13 sliders available for manipulating 13 parameters (also see Table 1) in the model. Note that in the Epidemic Curves (EPC) graph, the `Dead_(D/N)` button is in gray because this graph is switched off, though the total number of deaths are plotted (blue curve) in the Accumulated Totals (ACT) graph. Note that the left control roller for `Kappa` (KAP) is in green because the `Constant` mode (roller value 1) for this RAM has been selected rather than the `Default` (which would be in blue and the roller value would be 0), which is the `Adaptive Contact` mode expressed in Eq. 10 (also see Fig. 4).

***<u>Top ribbon</u>*** *(from left to right)*

##### Run length window

Type in the length of the run and then press ‘enter’ on your keyboard

##### S access window button (S On)

This window provides the user for a place to load a script written in Numerus Blackbox Programming Language (NBPL) or to write a de Novo script to control and make the current run (see Fig. 4).

##### RAM access button (Op On)

This will open the window that allows the user to access the different runtime alternative modules (RAMs) where options are available to use alternative functional descriptions or enter a ‘user-defined description’ of a key process in the model.

##### JavaScript window button (Op On)

The user can open a window and either load in a script or write a JavaScript to control a run using the `Airport codes` to access variables and functions (See Fig. 7)

##### dt window

This window only appears in the continuous time (and not in the stochastic) RAMP. It is size of the numerical integration interval used to solve the underlying system equations using a Runge-Kutta 4 algorithm.

##### Reset button

Use to clear graphs and data from previous run.

##### Single step button

Each time this button is pressed the simulation will progress by one time step.

##### Run button

After pressing reset to clear previous run data, press this button to initiate a new run.

##### Run progress window

In this window, the progress of the computation is monitored by counting down in time from the length of simulation to 0 (i.e., end of simulation).

***<u>Main window</u>*** *(from left to right and top to bottom)*

##### Parameter sliders Name (Airport Code, mathematical symbol)

`Baseline contact rate` (KP0, *κ*_0_) This parameter in Eq. 10 is the contact rate at the start of the epidemic (i.e., *κ*(0) = *κ*_0_).

`Contact rate scaling` (KPS, *P*_*κ*_) This parameter in Eq. 10 is the prevalence (proportion of infectious individuals) at which the baseline contact rate is reduced by a half (i.e., *κ*(*t*) = *κ*_0_/2 when *I*(*t*)/*N*(*t*) = *P*_*κ*_).

`Transmission parameter` (BET, *β*) This parameter, interpreted as the ‘force of infection’ appears in Eqs. 7 and 8, as seen in Eq. 12, when multiplied by the contact rate *κ*(*t*) essentially determines the probability of infection per contact.

`Recovery rate` (RIR, *ρ*_IR_) This is the flow rate from the infectious to the immune disease class: it also is the inverse the infectious period (see Eq. 1.2 in Box 1).

`Loss-of-immunity rate` (RRS, *ρ*_IR_ ≡ *ρ*_IV_) These are the rates at which individuals return from disease classes R and V (for simplicity assumed equal in our model) to become susceptible again in disease class S. Their inverse is the period immunity remains effective (see Eq. 1.2 in Box 1).

`Disease induced death rate` (RID, *ρ*_ID_) This parameter, referred to as the virulence parameter *α* by mathematical ecologists specifies the per capita prevalence rate at which infected individuals die over time.

`Treat` (TRT, *ρ*_treat_ ) This parameter is the rate at which infectious individuals are placed under treatment, provided the treatment capacity *T* ^max^ has not been exceeded (see Eq. 21 for details of how *ρ*_IT_ is defined in terms of *ρ*_treat_ and *T* ^max^). A key effect of treatment is to reduce the rate of contact of infectious individuals in class T compared with those in class I. In our model, we assume that individuals in class T no longer transmit pathogens to susceptible individuals, although in more complex models some transmission may occur, particularly to healthcare workers (but this is assumed to be negligible in our formulation).

`Treatment capacity` (TRC, *T*^max^) This parameter places a limit on how many individuals can be under treatment at any one time. When this level is reached, then new patients are limited by the rate at which individuals leave treatment because of death or recovery.

`Treatment discharge rate` (RTR, *ρ*_TR_) This is the rate at which individuals under treatment are released to join the immune class R.

`Death rate under treatment` (RTD, *ρ*_TD_) This is the rate at which individuals under treatment die from the disease. This rate may be greater or less than the disease-induced death rate for individuals in disease class I—the former when only the most severe cases are treated, and the latter when treatment is at least somewhat effective.

`Vaccination onset time` 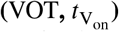 This is the time that the vaccination rollout program begins on the interval [0, *t*_fnl_] (i.e., it may be the start of the simulation or some way into the simulation).

`Vaccination regimens available` (VAM, *V* ^max^) This parameter sets an upper limit to the total number of vaccination regimens that can be administered (whether a regimen consists of one, two or more shots in a defined space of time, the number of regimens rather than shots are counted)

`Vaccination rate` (VAR, *v*) This rate only applies for 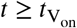 and as long as *V* ^max^ has not been reached.

**RAMs** `Code name, Descriptive name` (Airport code, mathematical symbol) in blue when default option is selected (roller value 0) and in green when an alternative value is selected (roller value 1 corresponds to constant values over the simulation interval, roller values > 1 will be user supplied options; text in red if the reset button needs to be toggled)

`Kappa, Adaptive Contact` (KAP, *κ*(*t*)) The `Default` (roller 0) for this expression, as given by Eq. 10, involves the baseline contact rate *κ*_0_, the contact rate scaling constant *P*_*κ*_ and the transmission parameter *β*. Alternate roller 1, labeled `Constant`, is the constant value *κ*_0_.

`Treat_Rate, Treatment Flow Rate` (RIT, *ρ*_IT_) The `Default` (roller 0) for this expression, as given by Eq. 21, involves the baseline treatment rate *ρ*_treat_ and the treatment capacity parameter *T* ^max^. Alternate roller 1, labeled `Constant`, is the constant value *ρ*_treat_.

`rSV, Vaccinate S` (RSV, *ρ*_SV_) The `Default` (roller 0) for this expression, as given by Eq. 23, involves the baseline treatment rate *v*, treatment onset time 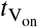 and the treatment regimens available parameter *V*^max^. Alternate roller 1, labeled `Constant`, is the constant value *v*.

`rRV, Vaccinate R` (RRV, *ρ*_RV_) The `Default` (roller 0) and alternative `Constant` (roller 0) as exactly the `same as Constant Vaccinate S`. This option has been provided in case the user wants to vaccinate disease class R in a different manner to disease class S. This would require the classes R and S can be distinguished by the healthcare establishement.

##### Graphs Name (Airport code)

`Epidemic curves` (EPC) This graph allows the user to plot the proportion of individuals in disease classes S, I, R, and D over the chosen simulation interval or to select one of these four variables to create a comparative graph over multiple runs.

`Accumulated totals` (ACT) This graph allows the user to plot the accumulating number of deaths (*D* (*t*)), vaccinated *V*^total^(*t*)) and treated (*T* ^total^(*t*)) individuals as time progress, or to select one of these three variables to create a comparative graph over multiple runs.

`Epidemic progress` (EPP) This graph allows the user to plot the effective reproductive rate (*R*_eff_(*t*)) or prevalence (*I*(*t*), repeat of second graph in EPC) over time, or to select one of these two variables to create a comparative graph over multiple runs.

#### 2.5.2. Stochastic SIRS+DTV RAMP dashboard

As with the Continuous-time SIRS+DTV RAMP, the latest available version of the Stochastic SIRS+DTV RAMP can be downloaded at our Numerus Model Builder Website where more information on using the RAMP is available. The dashboard of the Stochastic SIRS+DTV RAMP is the same as its continuous time counterpart, except the dt window is now replaced as follows:

##### Seed window

This window (center of top ribbon) anchors the sequence of pseudo random numbers used to in the simulation to a particular starting point: if this window is empty, then the starting point is random, otherwise it can be specified by a number ranging for 0 to 2^64^ − 1.

There are also one additional slider in the stochastic dashboard

##### Parameter sliders

`Name` (Airport Code, mathematical symbol)

`Popsize` (PSZ, *N*_0_) This parameter is the initial population size *N*_0_ that appears in Eq. 24, where the initial number in disease class *S* is *S*(0) = *N*_0_ − 1.

The reason why population size does not appear in the Continuous-time SIRS+DTV RAMP is that the solution to continuous time model Eq. 20 is scale free relative to the size of *N*_0_ (i.e. every equation in this model can be divided by *N*_0_ to obtain a system of equations on the time course of the proportions *S*/*N*_0_, *I*/*N*_0_, *R*/*N*_0_, *T* /*N*_0_, and *V* /*N*_0_, so that the equations are now independent of our choice of *N*_0_, provided the initial conditions are set to the corresponding set of fractional values. On the other hand, the variance associated with demographic stochasticity depends on absolute values (viz., the variance associate with multinomial sampling is inversely proportional to the square root of population size), which implies that the behavior of the initial outbreak is nearly (i.e., provided *S*(*t*)/*N*(*t*) remains very close to 1) independent of population size provided the transmission is purely frequency dependent (i.e, does not have a density dependent component to it).

Every run of the stochastic RAMP for a given set of parameter values will be slightly different, unless these runs are made with the same value inserted in the `Seed` slot in the middle of the top ribbon of the stochastic SIRS RAMP (Fig. 3). To make sure that the same seed is used at the start of a new run, the `Restart RNG on Reset` square below the `seed` window can be checked. To answer many questions, it is often likely to be more useful to make multiple runs and provide statistics related to quantities of interest, than to make and present a single run. This idea will be further developed in the Section 3.4. In terms of visual interest, repeated runs can be compared by selecting the `Comparison` mode slot above each graph, and making sure that all by one of the graphs that can be plotted with each graph (see lists below each graph in Fig. 2, which can be selected or deselected by clicking on the appropriate small circular button. In the two graphs in Fig. 3A. the proportion of immune individuals and number of infectious individuals are plotted for 10 comparative runs for set of values indicated in the slider windows and in Table 1 when not available for slider selection.

**Figure 3:**
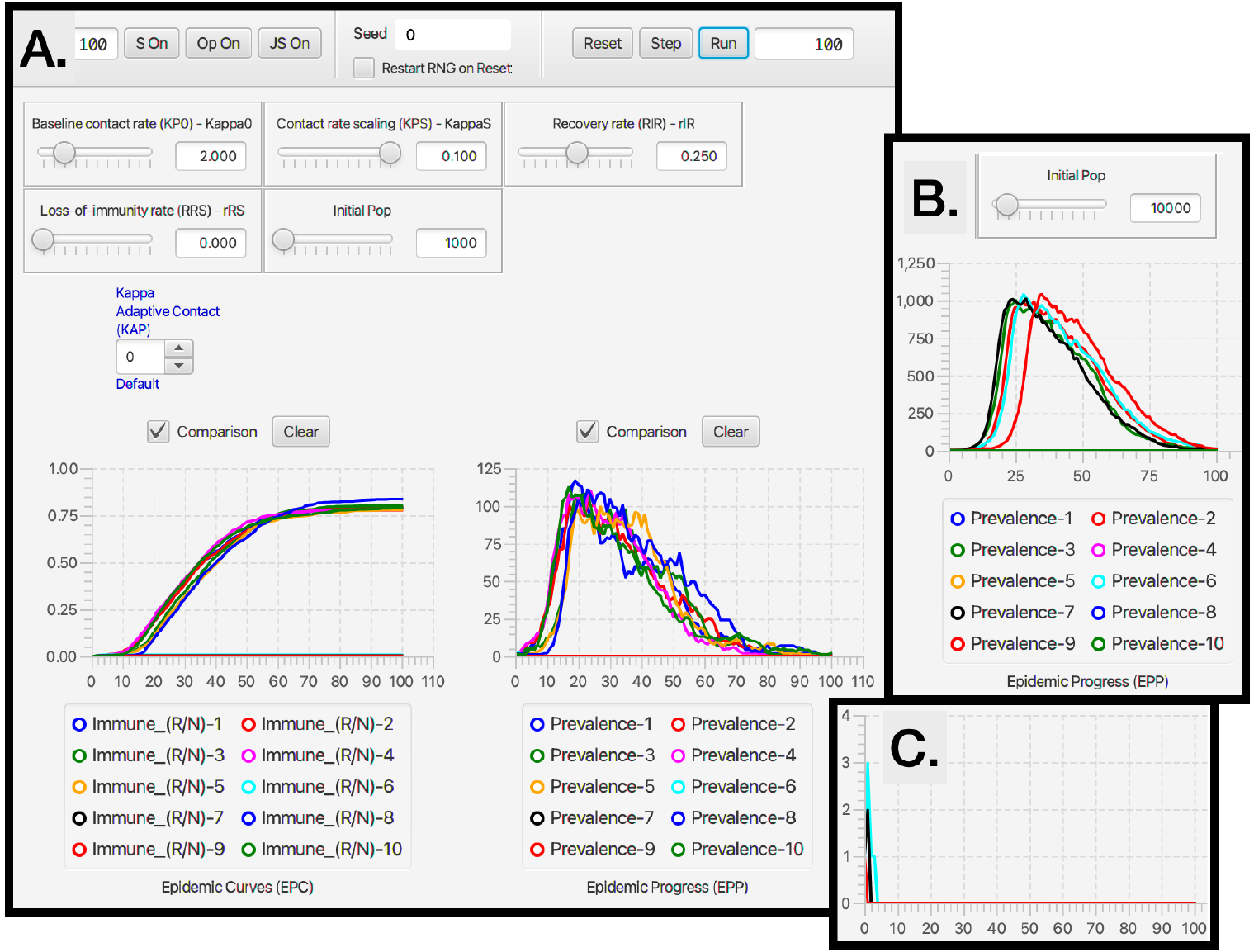
Ten comparative runs of the stochastic SIRS RAMP using the parameters values depicted by the sliders (or fixed according to values listed in Table 1). In panel **A**. the population size is 1000, in panel **B**. it is 10,000, while in panel **C**. a vertically magnified version of prevalences for the three runs that failed to take off in panel A (Runs 6, 7 and 9; such runs are expected, as discussed in the Results Section in the context of Eq. 25) are plotted on their own.

**Figure 4:**
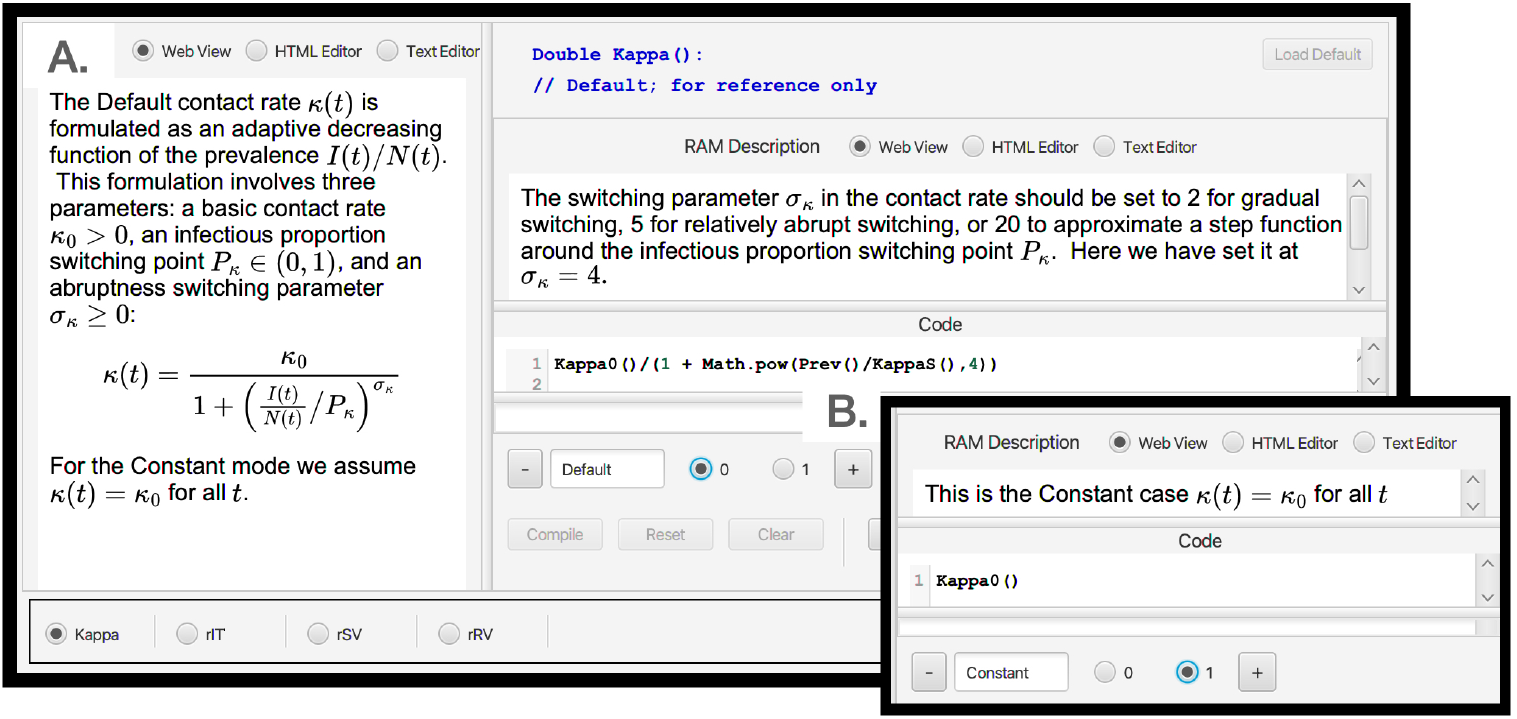
When the `Op On` button (second button from the left on the top strip of an SIRS+DTV RAMP, as depicted in Fig. 2, is selected the window depicted here opens (panel **A**.). At the bottom left hand side we see that we have four RAMs (runtime alterable modules) options pertaining to the formulations of the functions `Kappa` (*κ*(*t*)), `rIT` (*ρ*_IT_), `rSV` (*ρ*_SV_) and `rRV` (*ρ*_RV_). We also see that the `Kappa` RAM has been selected. The `Default` option (number 0) of this RAM is the adaptive form given by Eq. 10 (it appears in the window on the left of panel **A**., with some specific details pertaining to this mode provided in the window on the right of panel **A**.). Inset **B**. shows the alternate form (number 1) for *κ*(*t*), which is simply setting it equal to the constant *κ*_0_. The user may insert an additional alternate form for this RAM by selecting the ‘plus’ button, bottom middle of panel **B**., and then adding the desired from in the `Code` window on the left-hand side.

**Table 1.** Parameter values and ranges used in illustrative simulations

| Parameter | Symbol | Value/Range | Comment |
| --- | --- | --- | --- |
| Time unit | $t$ | daily | |
| Nominal pop size* | $N_0$ | $10^5 - 10^7$ | see Eq. 19 |
| Simulation interval | $[0, t_{\text{fin}}]$ | varies | as specified in text |
| Baseline contact rate | $\kappa_0$ | 0.2-8 per day | see Eq. 10 |
| Transmission parameter | $\beta$ | 0.3 | see Eq. 7 |
| Infectious period | $\frac{1}{\rho_{\text{IR}}}$ | 4 days | median time in I |
| Period of immunity | $\frac{1}{\rho_{\text{RS}}} = \frac{1}{\rho_{\text{VS}}}$ | 10-∞ days | mean residence time in R & V |
| Disease-induced mort. | $\rho_{\text{ID}}$ | 0-0.01 per day | aka 'virulence' |
| Behavioral switch | $\frac{I(t)}{N(t)} = P_{\kappa}$ | 0.001-∞ | $\infty \equiv$ no behavior (Eq. 10) |
| Switch abruptness | $\sigma_{\kappa}$ | 4 | see Eq. 10 |
| Treatment rate | $\rho_{\text{IT}}$ | 0-0.1 per day | starts from day 1 |
| Treatment capacity | $T^{\text{max}}$ | 0-10,000 per day | see Eq. 21 |
| Vaccination rate | $\rho_{\text{SV}} = v$ | 0-0.01 per day | starts at 'Vacc. on' |
| Vacc. on | $t_{\text{V on}}$ | day 0 to 180 | see Eq. 23 |
| Vaccination capacity | $V^{\text{max}}$ | 100,000 in total | see Eq. 23 |

\*In the continuous-time model the actual value of this parameter has no influence on the dynamics and so all numbers are relative to its value, which can be used to normalize results to proportions or percentages. In the stochastic model, the actual value affects the level of variability as discussed in Sections 2.5.2 and 3.4.

#### 2.5.3. Operation of Runtime Alternative Modules (RAMs)

The four RAMs all operate in the same manner. The `Default` modes (value 0 on roller selectors and text in blue) for the `Adaptive Contact`, `Treatment Flow Rate`, `Vaccinate S` and `Vaccinate R` modules are respectively coded by Eqs. 10, 21, 23 and, again, 23. Along with these a Constant alternative mode (value 1 on the roller selectors and text in green, though text is in red when still to be included by pressing the `reset` button on the top ribbon of the Dashboard) is available for each of these. Information on these alternatives is provided in the window opened by toggling the `Op On` button, second from the left on top ribbon of the dashboard (Fig. 2). This window (Fig. 4A.) allows the user to select the RAM of interest (bottom left hand list). Once selected (in Fig. 4A., for say the `Adaptive Contact` RAM, the `Default Kappa` option is shown) the window provides an option to select the `Default` RAM (left text inner window contains general information; the right top text inner window contains specific information pertaining to the particular alternative selected; and the bottom right inner window contains the coded expression that will be used). Panel B, Fig. 4B.) depicts selection of alternative 1 for the `Kappa` RAM along with the its specific information inner window and coding inner window. Next to the buttons to select the alternatives 0 and 1 is a “+” button (bottom of panel B, Fig. 4) that users can be use to add as many alternatives numbered 2, 3,… etc., as desired.

#### 2.5.4. Scripting windows

Two types of scripting windows are available. The `S On` and `JS On` buttons (first and third buttons on top ribbon in Fig. 2) can be used to write or load in scripts respectively written RPL (RAMP Programming Language; Fig. 4) or JavaScript. These scripts enable the user to control individual runs either by changing parameter values and RAM settings, or make multiple runs (Fig. 4) and plot these using the comparative window facility (Fig. 6). Accessing the window in which this script can be entered or loaded from a saved file is described in Fig. 4‘s legend, as is accessing the window containing a list of the RPL commands that can be used to write such scripts.

**Figure 5:**
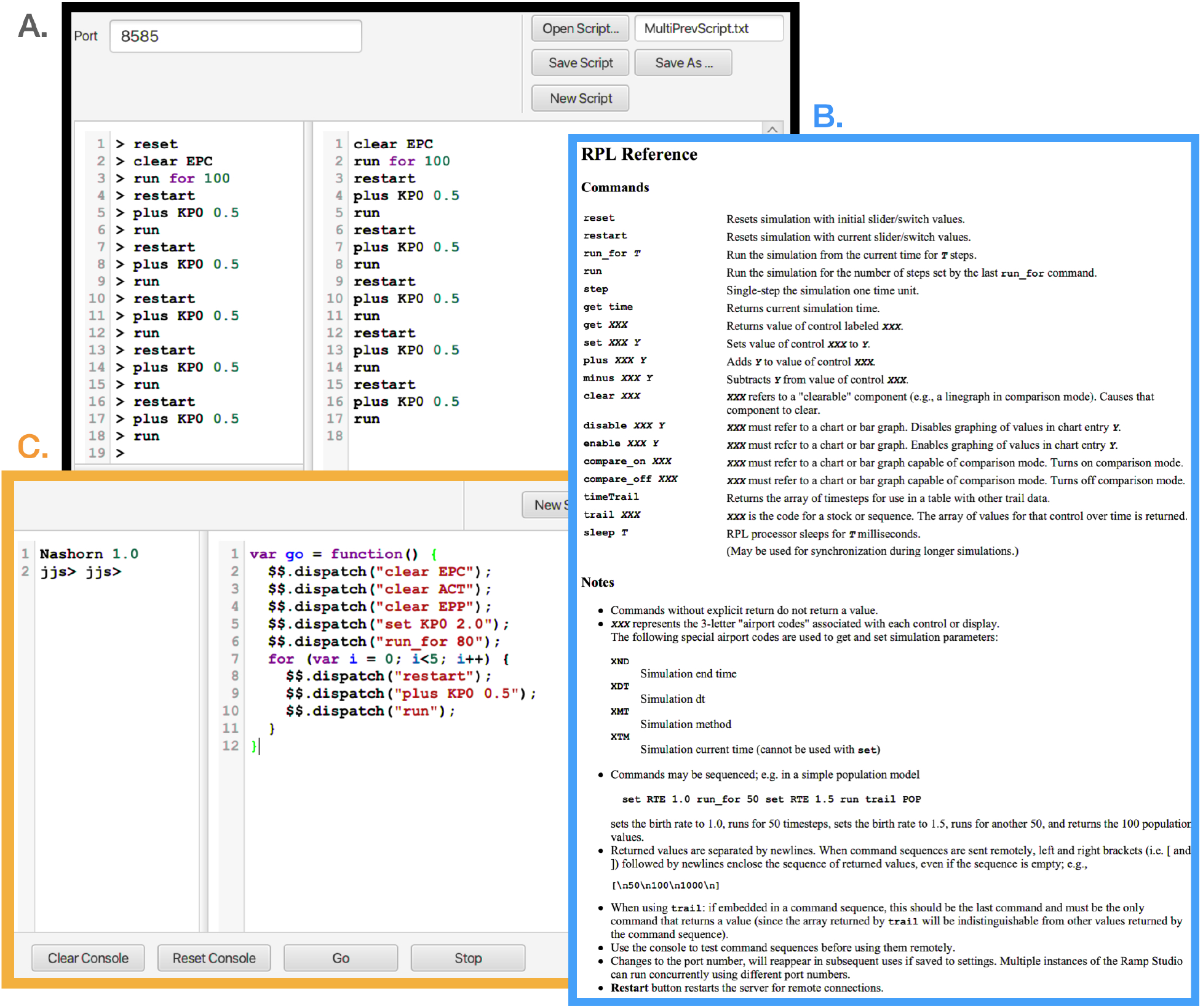
Here we depict three panels or windows associated with the scripting utilities of our RAMPs. **Panel A** (black bordered panel): When the `S On` button (first button from the left on the top strip of a RAMP, as depicted in Fig. 2) is selected this window opens. In the bottom left corner of this window is a `Command Reference` button (covered here by panel C), when pressed, opens panel B. **Panel B** (blue bordered panel): RPL Reference window, containing a list of commands that can be used to automate multiple runs or make changes to component values or modes during runs. The second column of text in the Script window (panel A.) contains 17 lines of instructions (saved as `MultiPrevScript.txt`, as seen in the top right hand window slot of the `S On` window) that codes the RAMP to make six repeated runs of the model, where in each of the runs the value of KP0 (*κ*_0_) is increased in value from 0.5, starting with the value specified in the model and described further in Fig. 6. **Panel C** (orange bordered panel): When the `JS On` button (third button from the left on the top strip of the RAMP, as depicted in Fig. 2) is selected this window opens. Here we see the JavaScript equivalent of the RPL script given in panel A.

**Figure 6:**
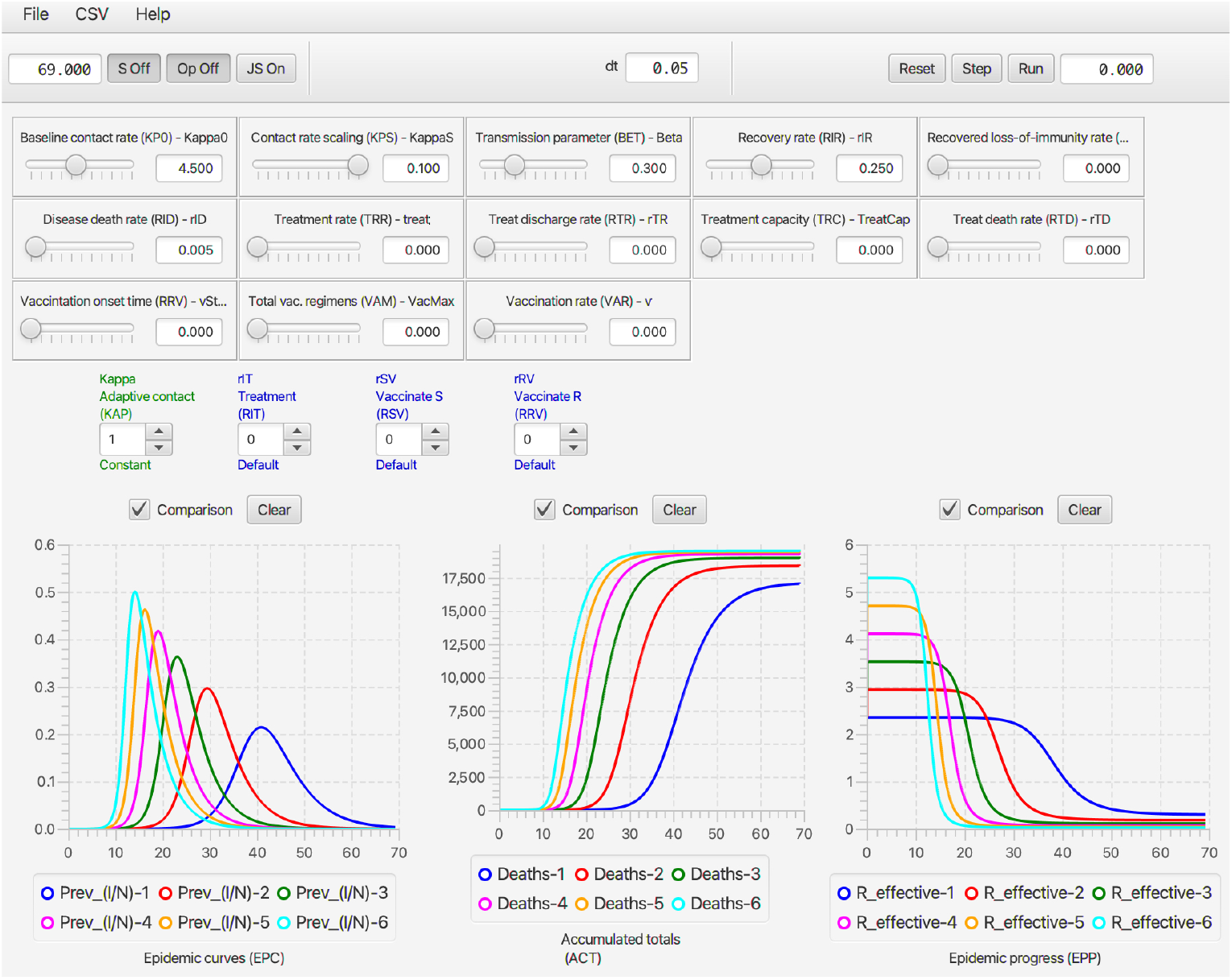
The script depicted in Fig. 5 is used to run the deterministic SIRS+DTV RAMP 6 times using the parameters values depicted by the sliders (or fixed according to values listed in Table 1). The KP0 (top left-hand) slider in this figure shows a value of 4.5, which implies the 6 runs ranged from a start value of 2.0 to 4.5 in increments of 0.5. Note that each of the three graphs available on the dashboard has only one its several possible options selected (available options can be seen below the graphs in Fig. 2) and the small rectangular comparison window above each graph has been selected to ensure that graphs generated during each of the 6 runs remain plotted once all runs are complete. The `c Clear` button above each graph allows earlier graphs to be erased when desired.

**Figure 7:**
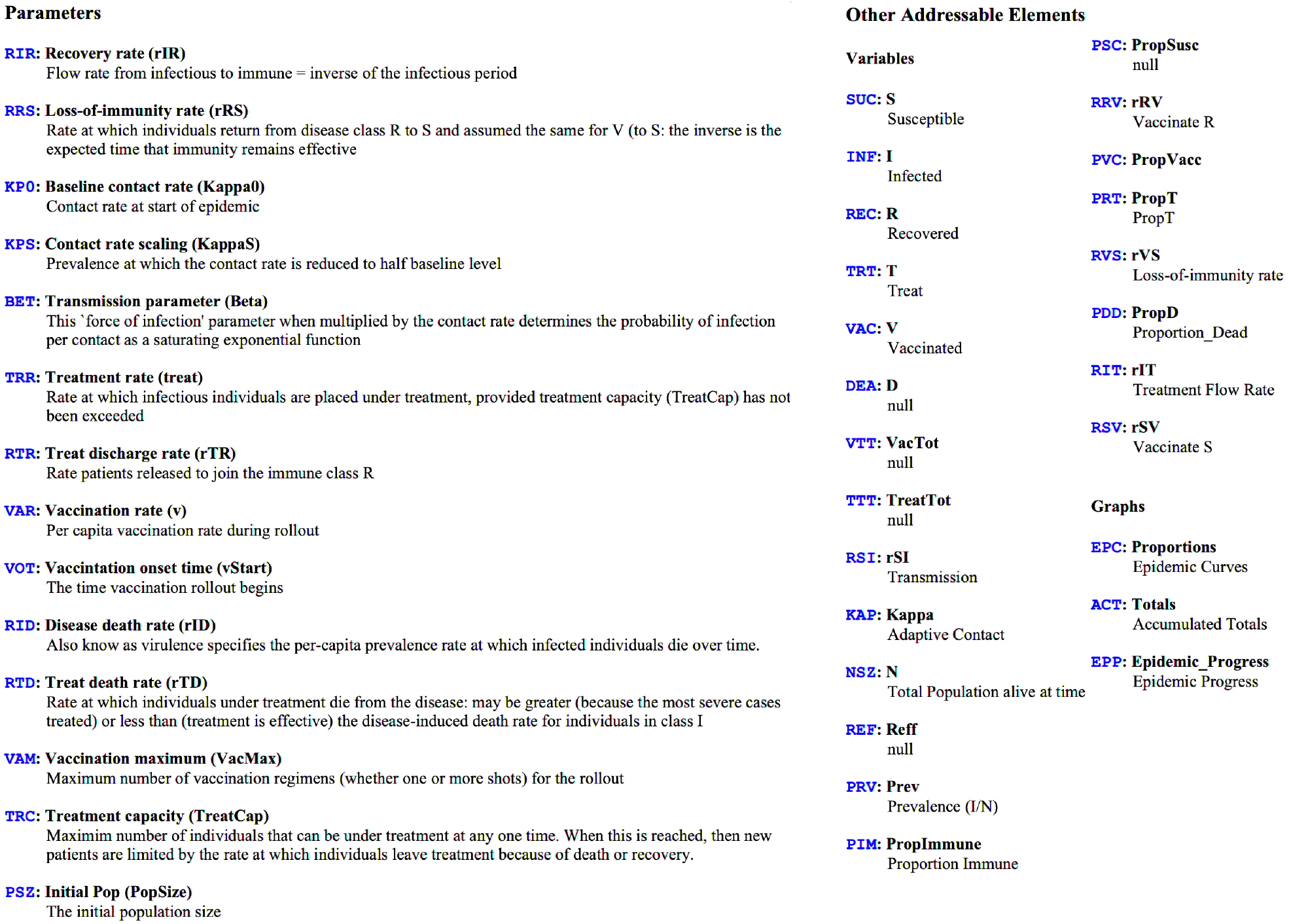
When the `Help` menu of RAMPs is selected information about the RAMP is provided. Here we see the information provided about the stochastic SIRS+DTV RAMP. This information includes the airport codes for the parameters, variables, and graphs, as well as descriptions associated with the sliders, which can also be accessed by mousing over the relevant sliders on the Dashboard.

#### 2.5.5. Help file and airport codes

When the Help menu of a RAMP is selected, information about the RAMP will appear together with a list of all the objects in the RAMP that can be referred to using airport codes (Fig. 7). In the parameter list on the left of Fig. 7), any parameter that has not been turned into a slider will have is *Fixed Value* listed in red. In our two SIRS RAMPs, though not in the our two SIRS+DTV RAMPs, the value of *β* (Airport code: `BET`) has been fixed 0.3 because only the product of *κβ* is relevant in determining the disease transmission rate *ρ*_SI_ for a particular prevalence level *I*(*t*)/*N*(*t*) (Eq. 7). The value of this product is thus manipulated using the *κ*_0_ slider, because *κ*_0_ scales the value of *κ* at time *t*, as expressed in Eq. 10.

#### 2.5.6. Saving the data from each run

A file location can be set up for the automated saving of CSV files (see Fig. 8 for details), which is particularly useful when the RAMP is used to make a series of runs, each differing in some sense (e.g., but changing a slider value and then rerunning the RAMP). The user can select what values should be saved using the `CSV Settings…` window. As indicated in Fig. 8, each file is saved with its own date stamp. Each CSV file is automatically headed with the set of parameter values and RAMP selections that were used to generate the data in the file.

**Figure 8:**
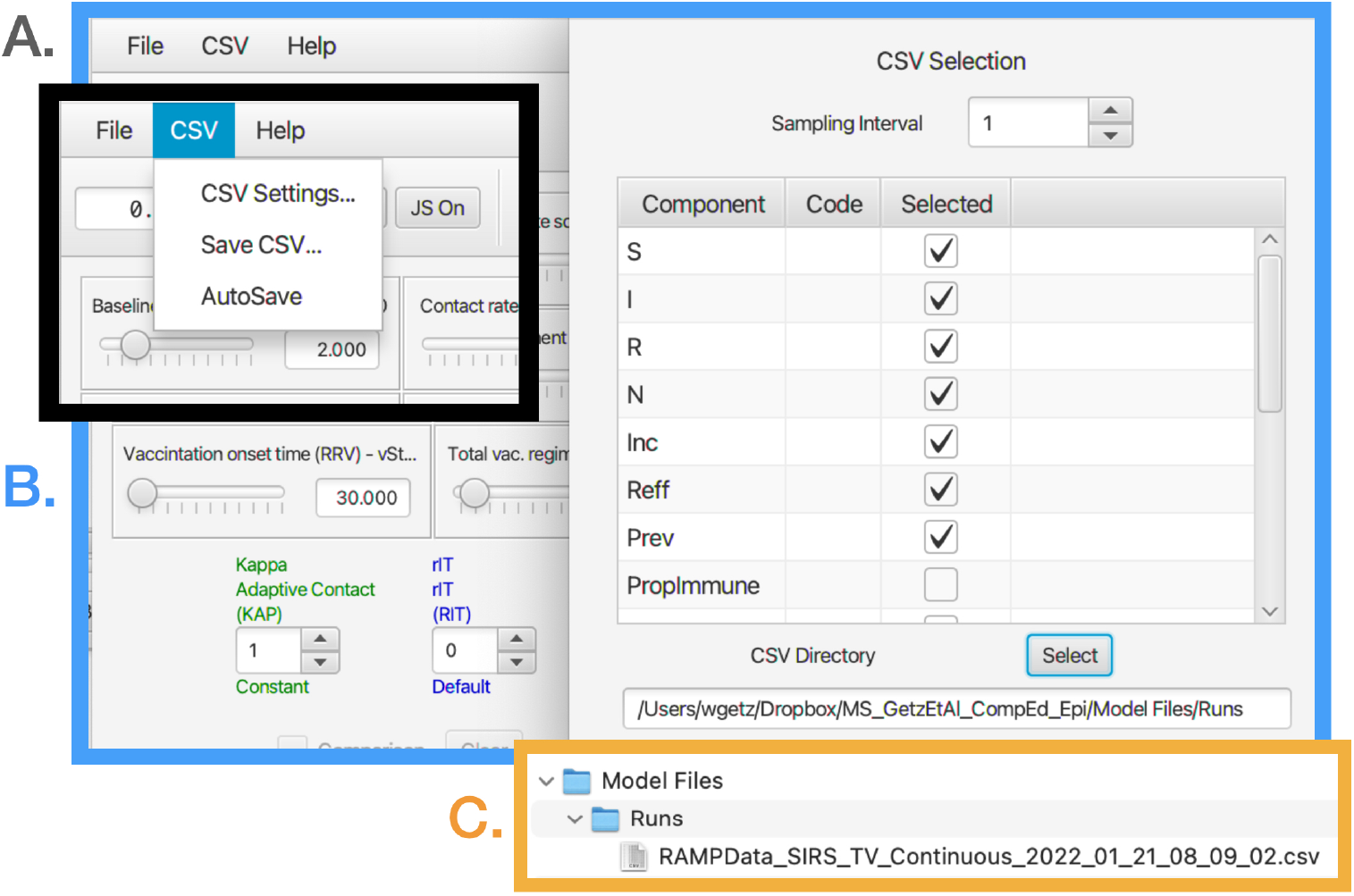
**A**. (black bordered panel) Each run can be saved at a user selected location that is set up using the pull down `CSV` menu on the top left-hand corner of the dashboard window (also see Fig. 2). **B**. (blue bordered panel) Once the `CSV Settings` option is selected a window opens that allows that user to select which disease states and computed time-series should be included in the CSV file which will be saved at the specified directory (example given here in the window at the bottom of the `CSV Selection` window. **C**. (orange bordered panel) Files saved in user specified directory are given a time and data stamp so that all files that are saved have a different name.

#### 2.5.7. Using RAMPs within the R statistical and data analysis platform

The exposition here applies to the use of Numerus Model Builder RAMPs as *virtual packages* in the implementation of R on the **RStudio** platform. The following steps should be followed to accomplish the match up between **RStudio** and **NMB Studio** (the Numerus Model Builder RAMP player) to implement the desired analyses.

1. Launch the RAMP to be used as an **RStudio** “virtual package” and either load the required parameter and RAM settings or choose these using the dashboard (Fig. 2).
2. Download the latest version of the `nmbR` package from the Numerus Studio site
3. Launch **NMB Studio** and install the package using the command `install.packages(“…/nmbR.tar.gz”, repos = NULL, type = “source”)` adding the appropriate directory.
4. Create the R code necessary to run the desired analysis, using the functions offered by the package, such as `nmbR$iterate_r` used in `ex1` in Fig. 9

**Figure 9:**
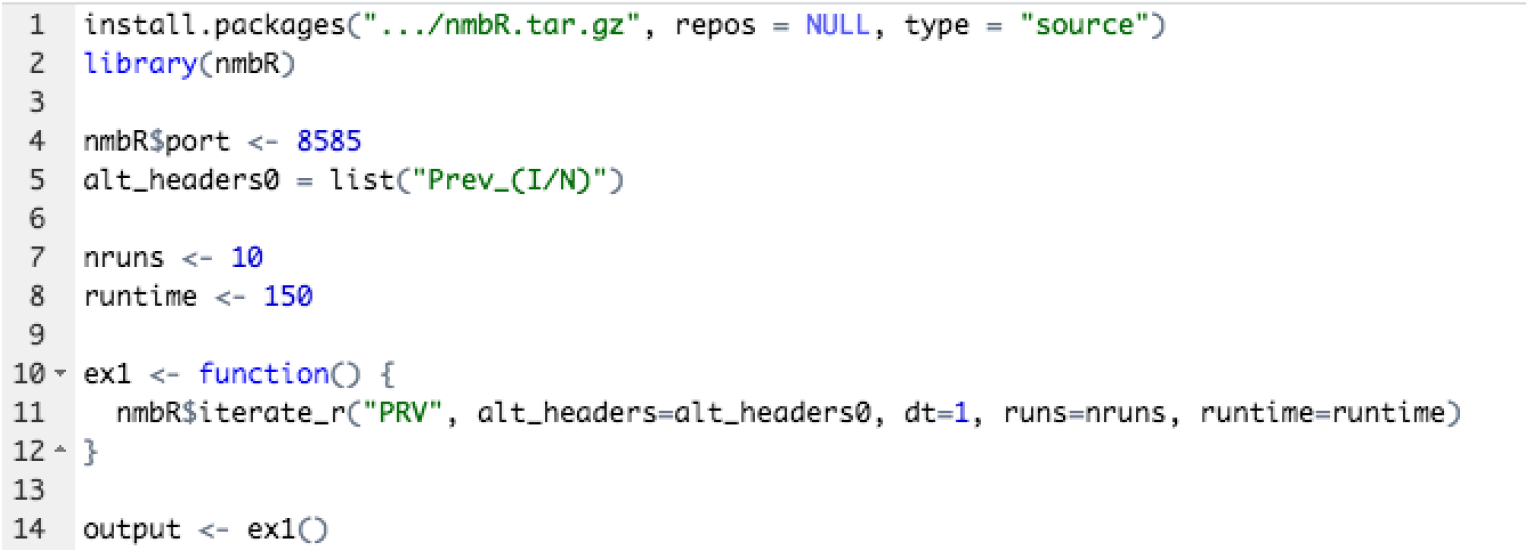
A partial capture of the **RStudio** window that is set up to run the `iterate_r` function of `nmbR package`. This package allows **R** code to utilize the RAMP currently running within **NMB Studio** as a ‘virtual’ **R** package.
5. Here is a list of the different functions defined in the package, while the user can define any other desired function using R code. For more information about the package functions, use the command `?nmbR` in the **RStudio** console.

`nmbR$dispatch`: to send a command string for processing by a running RAMP

`nmbR$iterate_r`: to iterate multiple runs, collecting in a list of per-run R data frames

`nmbR$iterate_v`: to iterate multiple runs, collecting in a list of per-variable R data frames

`nmbR$frame`: to retrieve the values generated in a RAMP simulation into an R data frame

`nmbR$graph`: to graph one or more time series from a RAMP simulation

## 3. Results: Illustration of Concepts

In this section, core epidemiological concepts and the efficacy of mitigating measures (treatment/isolation and vaccination) are illustrated through simulations, using parameter values and ranges of values listed in Table 1. These parameters are somewhat typical of a directly transmitted respiratory pathogen, such as COVID-19, but simplified by not including latent or asymptomatic disease classes Gao et al. (2021); Getz et al. (2021a), spatial structure (Riley, Eames, Isham, Mollison and Trapman, 2015), or age-class (Castillo-Chavez, Hethcote, Andreasen, Levin and Liu, 1989; Glasser et al., 2012) and other population structure effects (Getz and Lloyd-Smith, 2006).

In the six subsections that follow here, we present illustrative examples that each culminate in a take home message—i.e., six in total. We also provide numbered simulation exercises in Appendix A, were each exercise is numbered to link it to the material presented in each of the 6 subsections of this section.

### 3.1. Deterministic SIR epidemiological dynamics

In our first simulation, we demonstrate the basic features of an SIR (and, equivalently, an SEIR) epidemic process. The actual set of parameter values is not important provided they are selected to ensure that *R*_0_ > 1. Beyond this, different sets of parameters will result in epidemics that have the same primary features of an outbreak curve: a rise, a peak prevalence, followed by a collapse to zero prevalence with a proportion of susceptibles escaping infection. The only difference is 1.) how long it takes the epidemic to rise to its peak and collapse to almost zero (in idealized deterministic models the approach to zero is asymptotic; in stochastic models it reaches 0 in a finite time—see Section 3.4 for more details), 2.) the peak prevalence value itself, and 3.) the value of the proportion of individuals that escape infection.

For example, consider setting *κ*_0_ = 2 in the non-adaptive case (Eq. 9), *β* = 0.3, *ρ*_IR_ = 1/4 = 0.25, and *ρ*_ID_ = 0.05 (Table 3). In this case, it follows from Eq. 22 that 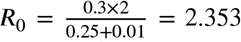. Since *R*_0_ > 1, we expect an outbreak to occur. As we see left most graph in Fig. 10, the curve *R*_eff_ (blue curve) starts out at *R*_eff_ (0) = *R*_0_ = 2.353. Despite *R*_0_ being almost 2 and half times larger than needed for an epidemic to occur, it takes time to make a visible impact in our plots, taking off around *t* = 26 (see prevalence curve in red, in the left graph).

**Figure 10:**
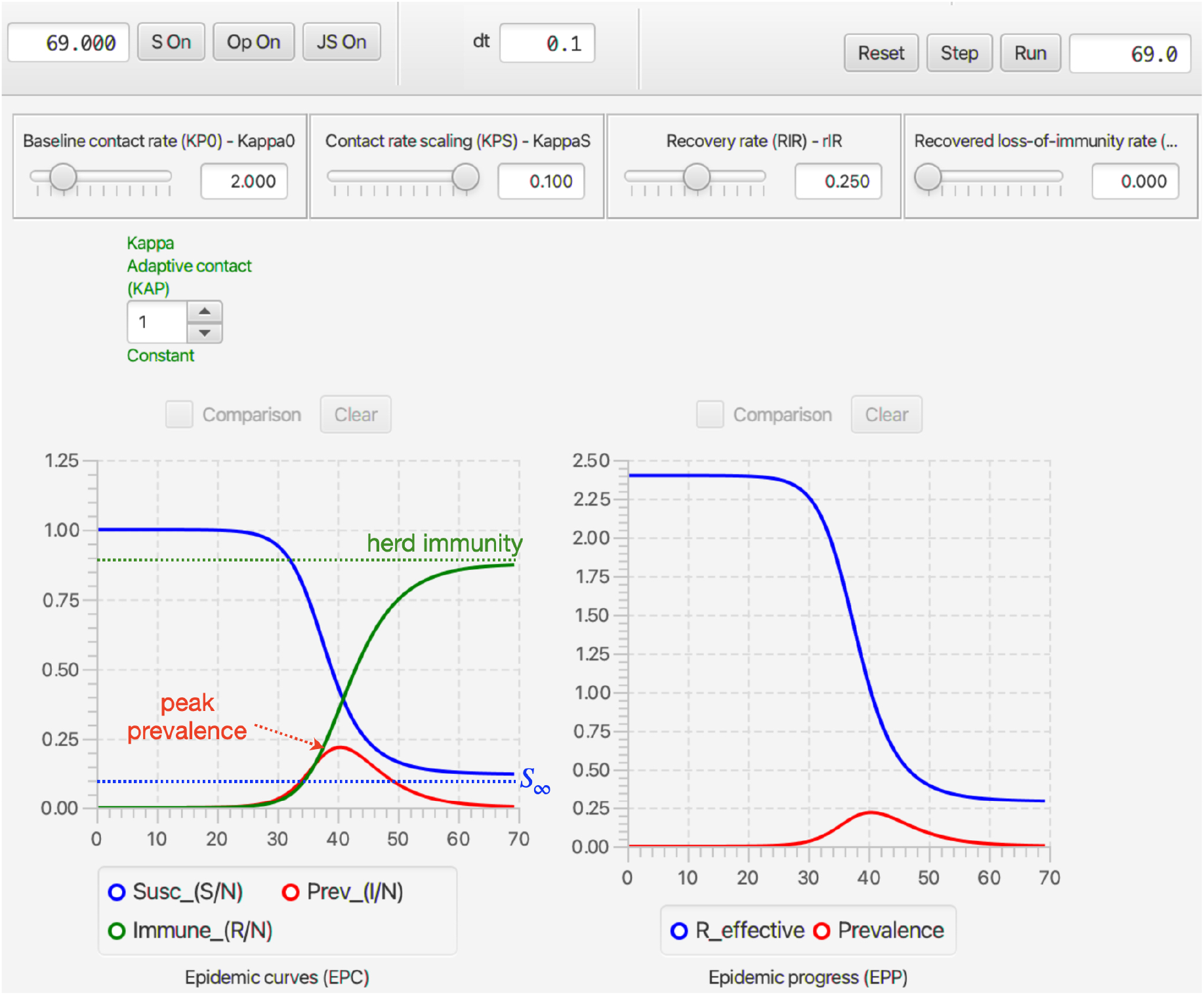
A view of the continuous-time deterministic SIRS RAMP dashboard after simulation of a basic set of epidemic SIR curves for the parameter values shown in the sliders, with an infectious period of 4 days, as listed in Table 1. Note the contact scaling value KappaS (aka behavioral switch, *P*_*κ*_ in Table 1) is moot because the `Kappa Adaptive Contact` RAM (green text in middle of panel) has been set to be `Constant` (roller setting is 1), while the setting for its `Default` mode would be 0 (also see Fig. 4). In this case *peak prevalence* is reached around 40 days, the *herd immunity* level is around 90% and the *asymptotic susceptibility* level *S*_∞_ is around 10%.

As the prevalence rises steeply after this take off, *R*_eff_ begins a concomitantly steep decline (blue curve, left graph), reaching a value of 1 around *t* = 40. At this same time, the prevalence curve (red, left graph) hits is maximum value of just under 25% and then rapidly declines to low levels around *t* = 60. Although deterministic models, as mentioned above, predict an asymptotic approach to 0 over infinite time, the last case will most likely disappear between 65-110 days (as discussed in Section 3.4), provided all of individuals in the R class (close to 90%, green curve right graph) remain fully immune (i.e., do not transition back to class S, as discussed in Section 3.2).

Of notable importance is that not all individuals succumb to infection: at time *t* = 100 days, the proportion of susceptibles is 0.124 (red curve, upper panel). The epidemic, though has run out of steam because most of the contacts between remaining infectious individuals I and others are with individuals from R who we have assumed cannot become reinfected, rather than from S. Further, at this time 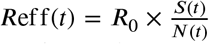 is much less than 1. In fact *R*_eff_ (70) ≈ 0.3. It is important to note that the epidemic does not shut down when *R*_eff_ (100) = 1 around *t* = 40 because it still has considerable momentum driving it down due to the prevalence at this point exceeding 20%.

#### Take Home Message 1

An SIR epidemic after rising to a peak falls to approach zero prevalence as the *herd immunity* level is approach, while the proportion of susceptible individuals approaches a non-zero value. Thus, once the epidemic has passed not all individuals will have succumbed to the disease.

### 3.2. Deterministic SIRS endemic dynamics

In an SIR model, individuals remain permanently in R and thus remain immune for the rest of their lives. This corresponds reasonably well to the case of immunity to measles (Naniche, 2009). If we take the effects of waning immunity into account by allowing individuals to transfer back to S, then we obtain an SIRS model. The mean rate of return from R to S can be obtained from empirical observations on the rates of reinfection of previously infected individuals as a function of time since last infection. When this rate is positive, the epidemic is not extinguished but reaches some endemic state that depends on this rate. The level of prevalence that the population finally settles into is inversely related to the mean period of time it takes for individuals in R to return to S (Fig. 11A.).

**Figure 11:**
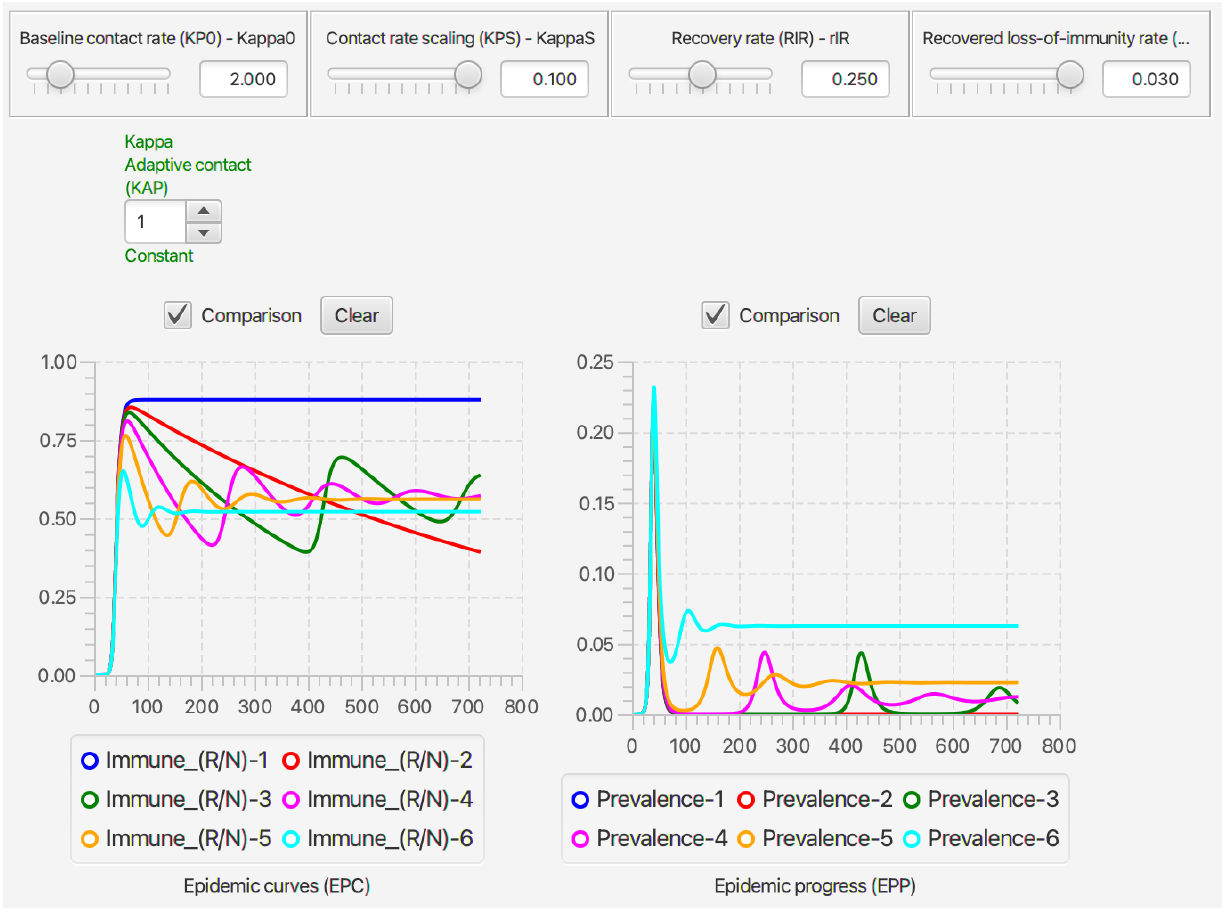
The level of immunity (*R*(*t*)/*N*, left graph) and prevalence (*I*(*t*)/*N*, right graph) are plotted over 720 days for a set of comparative simulations of the deterministic continuous-time SIRS RAMP to illustrate the phenomena of *endemicity* as a function of waning immunity. Plots 1-6 represent loss-of-immunity rates of *ρ*_RS_ = 0, 0.001, 0.002, 0.005, 0.01, and 0.03 respectively for individuals in class R. These corresponds to mean waiting (or residence) times in R (before returning to S) of ∞, 1000, 500, 200, 100, and 33.3 days respectively. Note that the value of the contact scaling rate KappaS= 0.100 is moot because the `Kappa adaptive contact RAM` (Green roller below sliders) has been selected to be the constant value *κ*_0_ = 2 (see Fig. 4 for more details).

#### Take Home Message 2

Levels of endemicity of a pathogen do not depend on *R*_0_, but on a given set of parameters that are inversely related to the waning period (i,e, mean residence time in R before returning to S).

### 3.3. Adaptive contact rates in the SIRS model

The trajectories depicted in Figs. 10 and 11 correspond to the case where the behavioral response is absent. An adaptive behavioral response can be implemented by setting the value *P*_*κ*_ in Eq. 10 to some finite positive value. For example *P*_*κ*_ = 0.1 implies that the basic contact rate *κ*_0_ is reduced by half (ie., to *κ*_0_/2) when prevalence is 10%. In Fig. 12B, we depict changes in the incidence curve for the basic set of parameter values in Table 3 for the SIRS model with mean recovery residence time is equal to 33.3 days rather than ∞ (note: the disease induced mortality rate in this case is 0).

**Figure 12:**
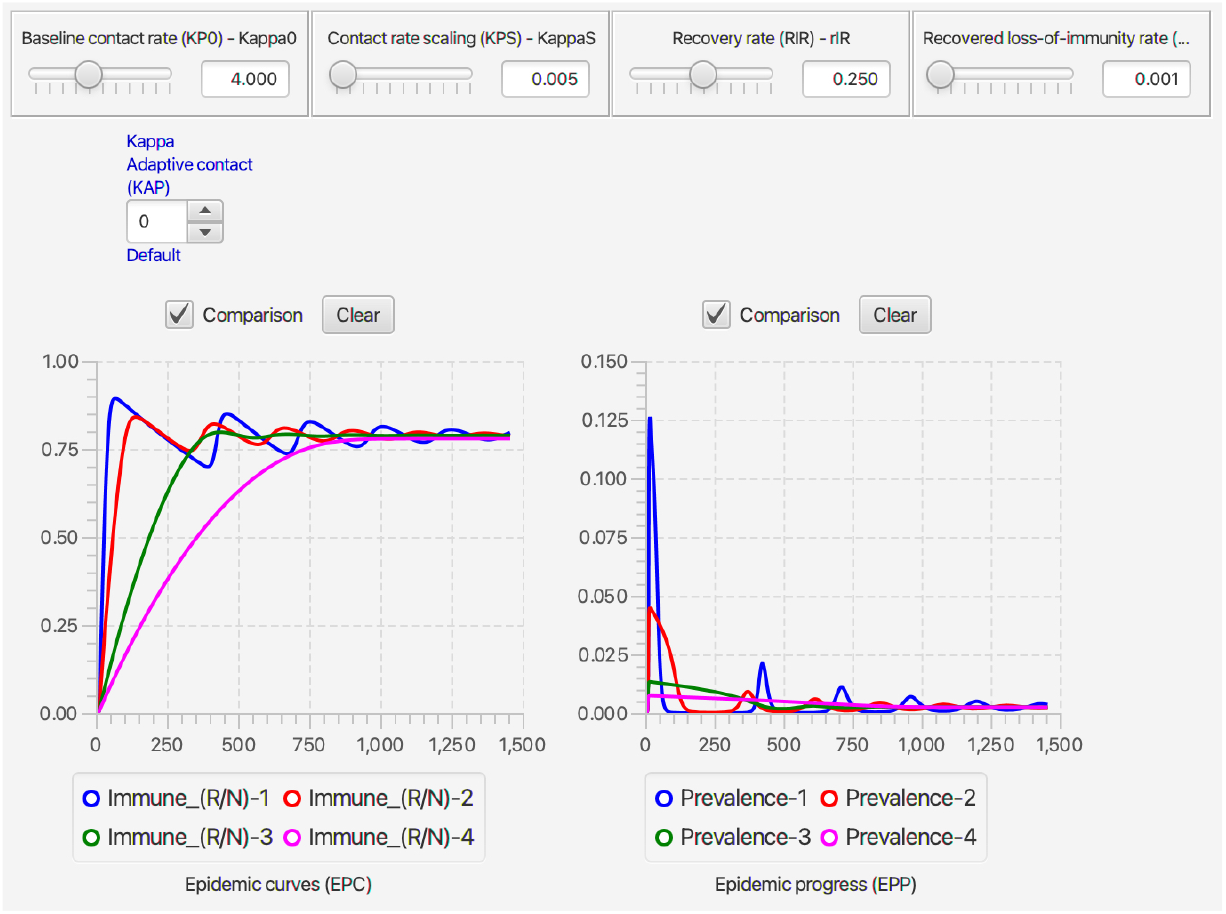
The level of immunity (*R*(*t*)/*N*, left graph) and prevalence (*I*(*t*)/*N*, right graph) are plotted over 1450 days for a set of comparative simulations of the deterministic continuous-time SIRS RAMP to illustrate the effect of *adaptive behavior* as a function of disease prevalence. Plots 1-6 represent scaling parameter values of `KappaS` (i.e., *P*_*κ*_ in Eq. 10) respectively equal to 0.1, 0.033, 0.01, and 0.001 Note that the `Kappa adaptive contact RAM` roller is now blue because the default 0 option applies, as discussed in Fig. 4.

#### Take Home Message 3

Adaptive social distancing can do much to dampen the peak response in a rising epidemic, but does little to help extinguish the epidemic or set the ultimate endemic levels.

### 3.4. Stochastic SIRS dynamics

Stochastic SIRS models of epidemics in large populations (e.g. tens of thousands or more) behave very much like deterministic SIRS models, except at the start and end of an epidemic when infectious class sizes are small (single digits). The properties of stochastic SIR dynamics have reported in considerable detail in text books and review papers Tuckwell and Williams (2007); Allen (2008); Britton (2010); Andersson and Britton (2012); Allen (2017), but here we will only focus on the most salient differences between stochastic and deterministic SIRS models.

Arguably the most important insight obtained from a stochastic model is that even when *R*_0_ >1, the introduction of an infectious individuals into a naïve population (i.e., with respect to the pathogen causing that disease or any related pathogens that may confer some cross-immunity in the population with respect to the pathogen causing the disease) does not automatically imply an outbreak of the associated disease will occur. In particular, it has been shown under certain assumptions, which we will refer to as the DH assumptions (see Diekmann and Heesterbeek 2000 for details of these assumptions), that the probability *p*^outbreak^ is related to *R*_0_ by the equation

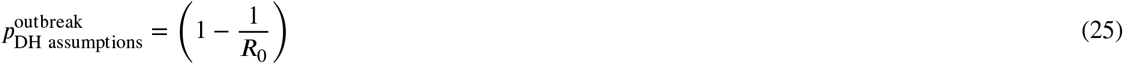

As an exercise, users of the Stochastic SIRS RAMP are asked to investigate how good an approximation Eq. 25 is for a SIRS epidemic modeled by Eq. 15.

A second important insight obtained from a stochastic model is that despite stochastic SIRS models predicting that all epidemics will ultimately become extinguished, endemic solutions persist for extended periods of time creating a so-called *quasi-stationary distribution* that depicts the distribution of prevalence values over time, given that the outbreak has past an initial *epidemic burn in phase* whose deterministic counterpart is the initial peak that we see in Fig. 11A before the equilibrium endemic level sets in (Britton, 2010; Riley et al., 2015; Allen, 2008).

#### Take Home Message 4

The value (1 – 1/*R*_0_) provides a reasonable, though biased estimate of the probability that an epidemic will start when an infectious individual is introduced into a naïve population.

### 3.5. Treatment

From a modeling point of treatment introduces considerable complexity into the model that may require additional structure to comprehensively evaluate its effects. For example, only overtly symptomatic cases are likely to be treated, and type of treatment will depend on severity of infection. In addition, we may ask: 1.) Does the introduction of treatment reduce mortality of cases in I that are not treated, because these are the milder cases? 2.) Does treatment reduce transmissibility overall due to possible isolation during treatment of the severer cases in I? 3.) How does treatment impact that mortality rate of those removed to class T (treated class)? And, 4.) Should the population be structured into a class of healthcare workers that are involved caring for individuals in class T, because these workers are now at risk of infection from individuals in T while the general population is not (e.g., see Lloyd-Smith et al. 2003). All this suggests that the most appropriate way to extend SIRS models to include a T class will be disease specific.

In terms of general analyses that can be undertaken when introducing treatment in the very simple manner depicted in Fig. 1 and expressed mathematically in Eq. 20, we can look at: 1.) the effect that introducing T has on reducing transmission in a homogeneous population because of its isolating components, and 2.) reductions to mortality through both treatment and the possible introduction of novel treatments partway through the pandemic.

The first case can be explored, for example, by reducing the transmission rate of individuals in T compared with hose in I. The second can be explored, for example, by assuming a relatively low disease-induced background mortality rate for all individuals left in I (perhaps even zero) and assuming that the disease-induced mortality rate of individuals in T is a decreasing (possibly step-wise) function of time as healthcare workers learn to deal with the new disease and pharmaceutical aids and methods of care improve steadily throughout the pandemic.

In Fig. 14 we compared two different scenarios that emphasize two aspects of treatment. To what extent is the capacity of the healthcare infrastructure going to be challenged by the peak number of individuals in treatment under the assumed treatment levels, and what are the expected number of individuals that will die from the disease over a specified period of time (in our cases 1000 days) under assumptions of how treatment will impact the mortality rate of those not in treatment, taking into account that those in treatment are the more severe cases anyway. The effect of scenario 2 compared with that of 1 is to reduce the peak number of individuals under treatment by around than 50% though due to different assumptions about the death rate under treatment leads to similar numbers of dead under both scenarios.

**Figure 13:**
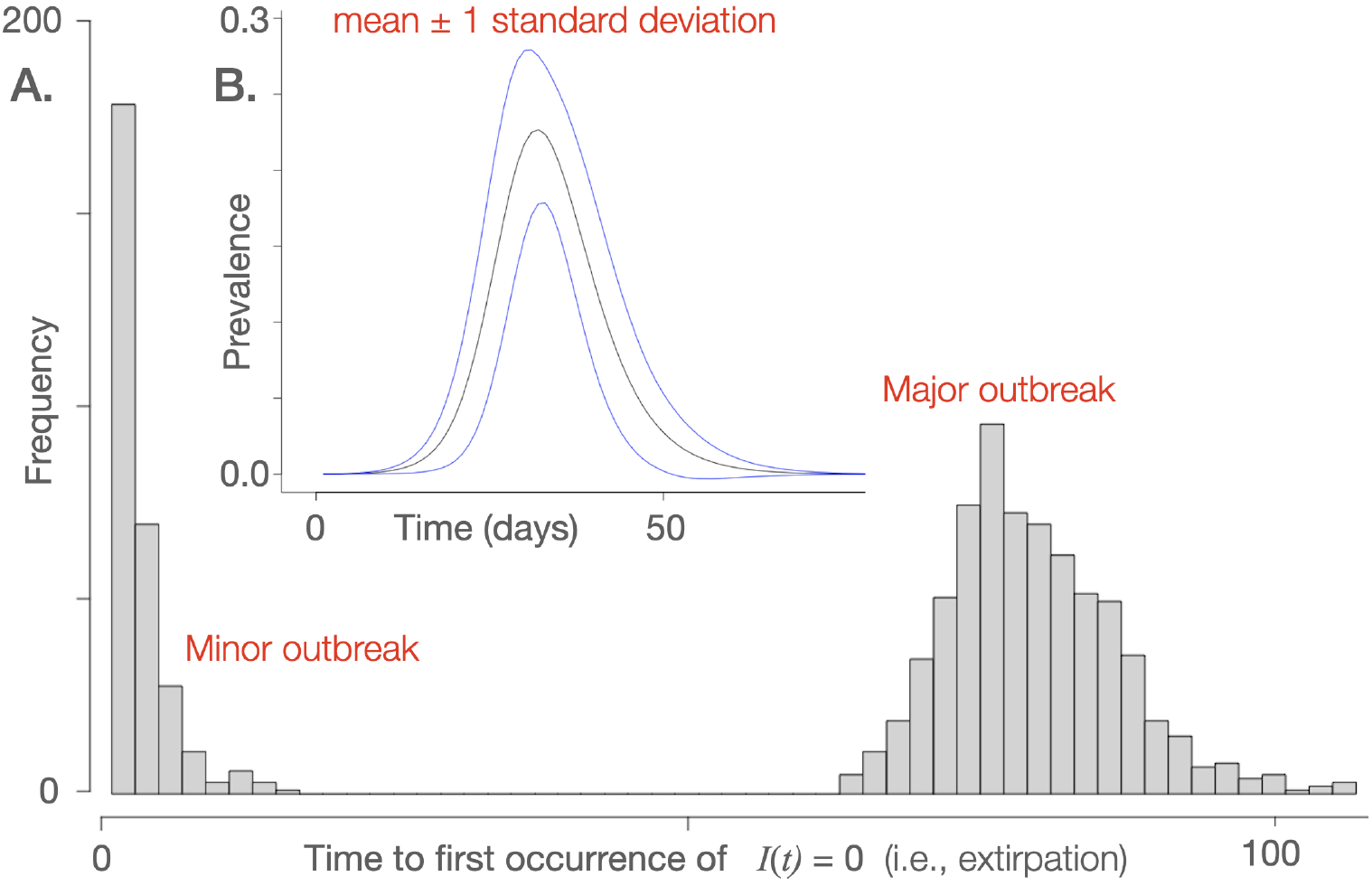
**A**. A histogram of the duration of outbreaks of an SIRS stochastic process obtained from 1000 simulations of the stochastic SIRS RAMP depicted in Fig. 3 embedded in an **RStudio** computational environment. This histogram clearly indicates a set of minor outbreak (301 simulations) and major outbreaks (699 simulations), where the former constitute a set of stuttering transmission chains that are extirpated before they can engage their possible exponential grow phase (Tritch and Allen, 2018). **B**. A plot of the mean and spread (± 1 standard deviation) of the prevalence over the 699 runs that constitute the major outbreak component of the 1000 simulations.

**Figure 14:**
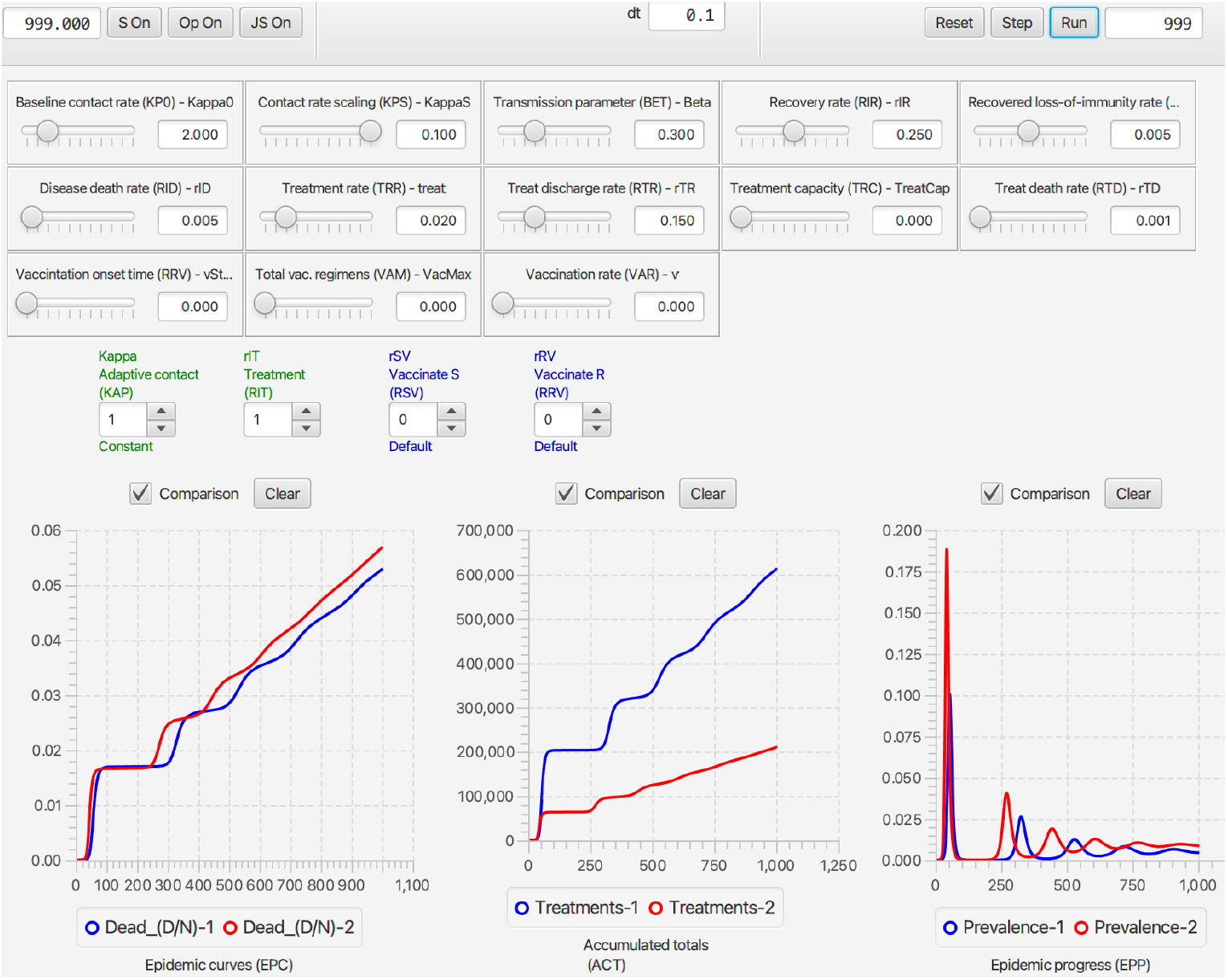
The per capita accumulating deaths (*D*(*t*)), the accumulating number of treatments (*T* ^total^(*t*)) and the prevalence (*I*(*t*)/*N*(*t*)), are plotted for two different constant treatment (i.e., *ρ*_IT_ RAM roller is set to 1) scenarios, with constant contact rate (*κ*_0_ = 2; `Adaptive contact` RAM set to 1) and no vaccination. In treatments 1 (blue curves) versus 2 (red curves) we have the treatment rates are *ρ*_IT_ = 0.1 versus 0.02 (10% versus 2%). The parameters used in treatment scenario 2 are shown in the sliders here. In scenario 1, we assumed that the disease induced mortality rate is the same for individuals, whether they remain in I or move T, which implicitly assumes that treatment is taking the more severe cases and reducing the mortality of these case to same background rate of 0.5% for those untreated. In scenario 2, we assumed that treatment is more effective than in treatment 1, reducing the mortality rate of those under treatment to 0.1%.

#### Take Home Message 5

The incorporation of treatment into an SIRS models has several critical aspects to it that may require alteration of the SIRS+DTV model to assess the effects involved.

### 3.6. Vaccination

Vaccination rollout programs are limited by the willingness of the population to participate, the healthcare facilities available to vaccinate individuals at a particular rate, and the number of regimes that will be available over the rollout implementation period. A recent exposition by an ISPOR (International Society for Pharmacoeconomics and Outcomes Research) task force recommended that economic analyses undertaken to implement new vaccination programs should consider the following four components (Mauskopf, Standaert, Connolly, Culyer, Garrison, Hutubessy, Jit, Pitman, Revill and Severens, 2018): “ 1.) uptake rate in the target population; 2.) vaccination program’s impact on disease cases in the population over time using a dynamic transmission epidemiologic model; 3.) vaccination program implementation and operating costs; 4.) and the changes in costs and health outcomes of the target disease(s).”

The RAMPs that describe here are meant to help implement component 2. of the above recommendation. By way of illustration of this component—ignoring components 1, 3 and 4 which would need to be brought into a more complete analysis at some point— we compare two vaccination programs: one starting on day 100 and another at 250 days, where both are constrained by a program rollout rate of vaccination 0.3% of all not currently or previously infected individuals per day (i.e. *ρ*_SV_ = *ρ*_RV_ = 0.003), with a maximum of one million regimens (irrespective of the number of shots associated with each regimen) available for the analysis.

The results of simulating the above two scenarios are illustrate in Fig. 15. The primary difference in the two scenarios is that the earlier start to the program helps suppress the second prevalence peak that occurs over days 200-300, and postponing it to a lesser peak over days 300-400. The early start does lead to a reduced proportion of deaths over days 200-700, but ultimately the advantages of the earlier start are nullified due to the earlier start program running out of vaccination regimens more than 100 days ahead of the later starting scenario.

**Figure 15:**
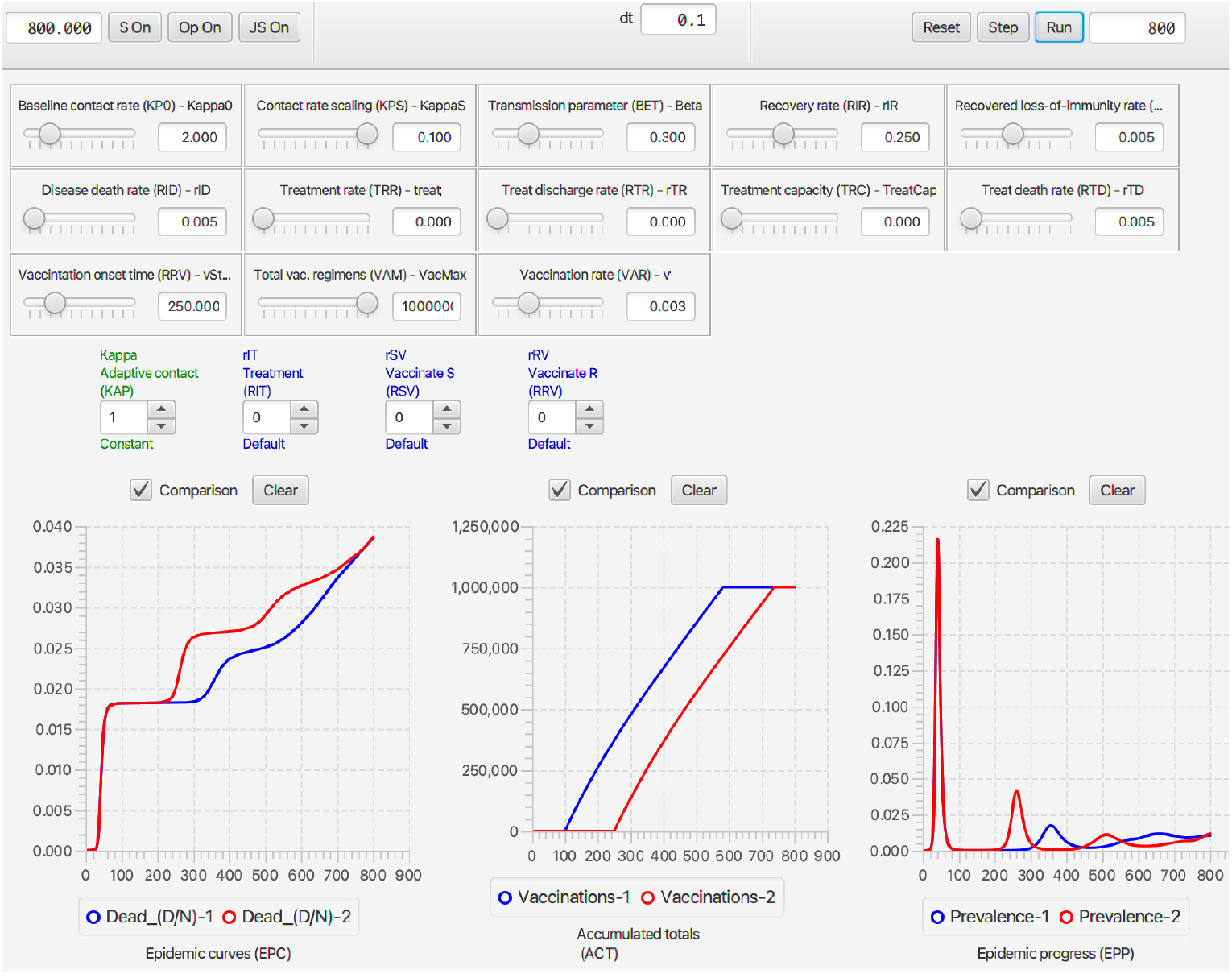
The prevalence (I), number of individuals currently immune through vaccination (V), and the number that have died are plotted for the two contrasting cases. In Vaccination Scenario 1 (blue curves) versus 2 (red curves) we have vaccination rates *ρ*_SV_ = *ρ*_RV_ = 0.003 (0.3% of individuals in these classes are vaccination each day) starting on day 100 for scenario 1 versus day 250 for scenario 2. Vaccinations are continued until one million regimens (whether done with a single or double shot is not relevant) have been delivered. Since the red curve initially obscures the blue curve, both scenarios are the same until day 100 (i.e., the initial red peak prevalence applies to both).

#### Take Home Message 6

Vaccination policies with limited regimens available over some fixed planning period may need to consider trade offs between peak patient loads that may stretch healthcare resources and mortality rates over the periods in question.

## 4. Discussion

Some of the most challenging global issues of our time—climate change, habitat and diversity loss, emerging diseases, and food security—cannot be mitigated effectively without an adequate understanding of how dynamical systems unfold in space and time. Notions of how best to describe the systems involved and measure their variables and rates of change are central to uncovering the spatiotemporal characteristics of these systems. Concepts needed to understand these characteristics include: the impact of time constants on exponential growth; how interacting variables, adaptive behavior and time delays may induced oscillatory patterns; and, the probabilities of invasion, persistence, and extinction of salient species as a function of various rates and process descriptions. Uncovering these characteristics requires some level of mathematical sophistication (e.g., mastery of the fundamentals of calculus) and dynamical systems modeling.

In the context of emerging disease and associated epidemiological processes, public health policy decisions often need to be made by health policy professionals, politicians, and civil servants. Many of these individuals do not have the appropriate mathematical training and modeling experience to undertake the kinds of quantitative analyses needed to make the soundest health policy decisions. Given how consequential these decisions may be with regard to the health of individuals and the economic well being of society, it is important that tools be developed to assist those involved in formulating or implementing health policy to understand the basic concepts involved. This is where simulation modeling tools come to the fore as instructional aids in teaching these concepts to individuals who lack the mathematical skills to build and analyze epidemiological models themselves.

In terms of tools to aid instruction in epidemiological dynamics, several groups have developed course notes and scripts to run SIR and moderately extended SIR models using the R or MATLAB software platforms (e.g., Mathematics Department at the University of Nebraska, Lincoln) or even spreadsheets (Ledder and Homp, 2020). These tools are not stand alone applications programs and require that students have a basic familiarity with the software coding platforms in question. Some educationally-oriented, web-based SIR modeling applications exist (Getz, Salter and Sippl-Swezey, 2015b), as well as stand-alone applications programs (Getz et al., 2018b). The utility of all these tools is limited by the relative simplicity of the models (e.g., they do not include treatment or vaccination options, and ignore the importance of adaptive contact behavior) and they do not have the flexibility of our RAMP technology to alter functional descriptions without needing to alter the underlying program codes.

Use of our RAMPs requires no coding or mathematical modeling skills, unless the user wants to write very simple scripts using our BPL or JAVA, or wants to use our RAMPs in conjunction with **RStudio**. It is important, though, that students are exposed to the primary assumptions underlying the construction of SIR models and their first line of extensions, as we provide in subsection 1.2.1. These assumptions provide insights into the limitation of the models and the actual complexity of the processes involved. The key to modeling is to capture the essential process involved and to understand the next level of processes that are being ignored by the models. This understanding facilitates assessment of the adequacy of the models as tools for making predictions and designing interventions (Larsen, Eppinga, Passalacqua, Getz, Rose and Liang, 2016; Getz et al., 2018a; Getz, Salter and Mgbara, 2019).

## 5. Conclusion

The RAMPs and RAMP player (**NMB Studio**) described in this paper, downloadable at the Numerus Model Builder Website, provide a set of tools that mathematically unsophisticated students can use to come to grips with the most important dynamically systems concepts associated with the outbreak of communicable diseases of epidemic and even pandemic proportions. The installation of these RAMPs on laptop and desktop computers is as easy as clicking a button to locate the download site and install the application. All the information needed to run the RAMPs is provided with the RAMP itself in the form of information in windows, with more substantial details contained in this paper and its supplementary information files. In closing we stress that our RAMPs and associated documents are meant to augment existing course material rather than replace such material, because the history, etiological and other important information associated with specific diseases (COVID, influenza, ebola, measles, tuberculosis, HIV, etc.), classes and types of disease (respiratory, hemorrhagic, bacterial, fungal, etc.) and modes of transmission (airborne, waterborne, vectored, soilborne, bodily fluid exchange, etc.) are not covered in this paper.

## Data Availability

All data produced in the present work are contained in the manuscript

https://www.numerusinc.com/studio

## CRediT authorship contribution statement

**Wayne M Getz:** Developed the models, generated the examples and figures, ran RAMP simulations, and wrote the manuscript with contributions and editing from the other authors.. **Richard Salter:** Developed and coded all the software for the two applications programs: NMB Designer and NMB Studio. The former can run be used to build and run models, as well as generate Numerus Model Builder (NMB) runtime alterable model platforms (RAMPs) as HTML files that can then be read and implemented by the latter as a stand alone application.. **Ludovica Luisa Vissat:** Was involved with the R implementation, ran R examples, checked all mathematical and RAMP coding components, and helped edit the final text..

## Appendices

### A. Simulation Exercises

The number of these exercises correspond to the material in the subsection numbers of Results Sections 3.1-3.6

#### Exercise 1

By varying the value of *κ* between 1 and 8, with other parameter values as Fig. 3, use the SIRS RAMP to:

1.1. explore the relationship between the time *t*^*^ and value *I*(*t*^*^)/*N*(*t*^*^) of maximum prevalence
1.2. explore the relationship between the time *t*^*^ of maximum prevalence and the time *t*_1_ when prevalence first rises above 0.1%
1.3 explore the relationship between the time *t*^*^ of maximum prevalence and the time *t*_2_ − *t*_1_ it takes for prevalence to drop below 0.1% at time *t*_2_, and note the value of *R*_eff_(*t*_2_) in each case
1.4 plot the proportion of individuals that escape infection in each case as a function of your computed value for *R*_0_ as a function of the selected value for *κ*.

#### Exercise 2

Plot (either as a series of 1-D curves or a 2-D surface) the relationship between the ratio of peak to endemic prevalence as a function of *R*_0_ (through manipulation of the value *κ*_0_) and the mean residence time 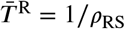

#### Exercise 3

By varying the strength of adaptive behavior through changes in the prevalence values of the behavioral switch parameter *P*_*κ*_ estimate the period of endemic oscillations that dampen over a five-year simulation interval as a function the values of *P*_*κ*_ and the mean waiting time 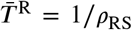 for combinations of these parameters where dampened oscillatory behavior is evident. (Note: this will require identification of local maxima in the prevalence curve over the five year simulation interval and computing the average time between consecutive local maxima over the five year period for each of selected pair of parameter values.)

#### Exercise 4

4.1. By varying the value of *κ*_0_ between 1 and 8, with other parameter values as in the stochastic SIRS RAMP depicted in Fig. 3, construct a histogram of times at which the prevalence *I*(*t*) becomes 0, as shown in Fig. 13. Note for each value *κ*_0_ at least 1000 runs should be made, as depicted in Fig. 13.
4.2. From from the runs made Ex. 4.1, plot the mean ± proportion (on the vertical axis) of runs in the major outbreak component and its standard deviation (assuming a binomial distribution for the mean and variance) as a function of the value of *R*_0_ (horizontal axis) that corresponds to the eight values of *κ* involved. Over lay this with a plot of Eq. 25 and comment on the fit.
4.3 (Advanced exercise) For *κ*_0_ = 4.0 and *ρ*_RS_ = 0.03, with other parameter values as in the stochastic SIRS RAMP depicted in Fig. 3, carry out 1000 simulations over the interval of time [0, 1200]. Select at random 10 of the simulations that constitutes a major outbreak (you might as well take the first 10 since the are randomly organized themselves), and plot a histogram of the 1000 values between *t* = 201 and *t* = 1200 for each of these 10 simulations. Compare normalized versus of these 10 histograms among themselves (i.e., reduce bins from numbers to proportions so the area under the histogram is 1—i.e., it is an empirical probability distribution). Also compare this histograms to normalized histograms of the values across all simulations that are part of the major outbreak at time *t* = 1200. Repeat this exercise for all values at time *t* = 1000. What do you notice. What does this tell you about the *ergodicity* of the SIRS stochastic model

#### Exercise 5

Come up with creative ways to explore the complexities of including treatment as a consequence of the rate at which individuals are treated, limitations on the number of individuals that can be treated at anyone time, and assumed effects of treatment. These effects include reducing mortality in the population as a whole and making assumptions about mortality rates for individuals that are treated.

#### Exercise 6

Come up with creative ways to explore the complexities of vaccination programs as a consequence of starting dates, vaccination rates, and limitations on the number of regimens available—either in absolute times or in monthly tranches.

### B. Boxcar Models

The exponential transfer distribution of Eq. 1.1 in Box 1 implies a maximum transfer rate of 1/*ρ* at *t* = 0 of a flow through disease class X. This is unrealistic since one would expect the maximum transfer rate not to be the moment of entering *X* but rather around the some mean period of time in X. This can be remedied by subdividing X into *K* sub-compartments x_*k*_, *k* = 1, …, *K* each of which is traversed at a rate *Kρ*. In this case we obtain a *boxcar transfer process through disease class X*, modeled by the following system of equations

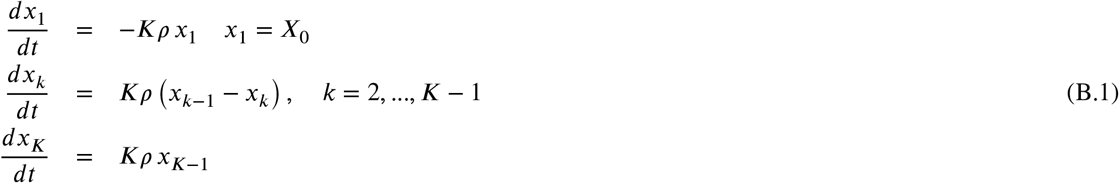

The solution X(t) to this systems of equation is known to be 1 minus the Erlang distribution multiplied by *X*_0_. Specifically,

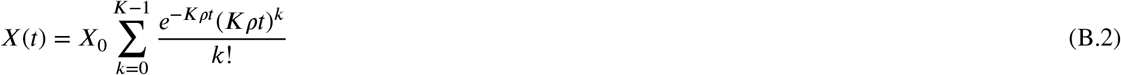

This also implies in the context of individuals passing through X—that is, through all *K* boxcars that constitute X—that

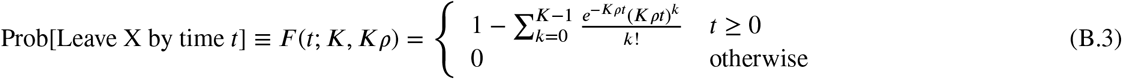

with corresponding Erlang probability density function (shape parameter *K*, scale parameter *Kρ*

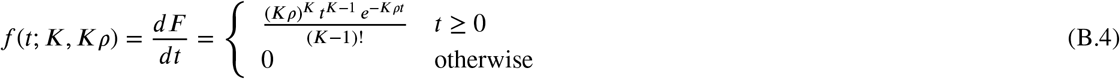

Thus, by computing 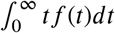, the mean time 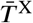 each individual spends in class X is

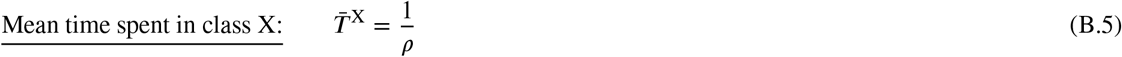

The mode is no longer at 0, but now is somewhat below the mean with a value 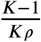. The variance of this distribution is 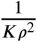 and as *K* → ∞, all individuals spend the same amount of time 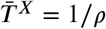 in X.

### C. Waiting Times in SIRS ABMs

Suppose the per capita outflow rate of individuals from disease class *X* is an increasing function of how long these individuals have been in X. We investigate the consequences of such an assumption by considering the exit distribution of *X*_0_ individuals who entered disease class X at time 0 under the assumption that their per capita rate of outflow from X is given by the function *ρ*(*t*) = *ρ*_0_*t*^*k*^, for constants *ρ*_0_ > 0 and *k ≥* 0. This process can be described by the differential equation (compare with equations in Box 1)

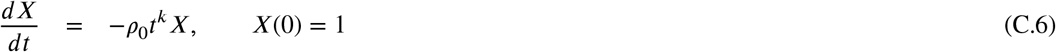

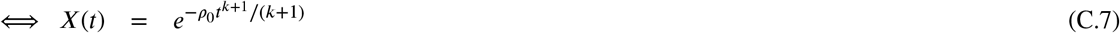

This also implies that

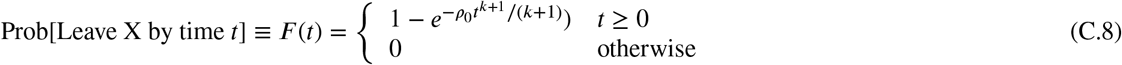

with corresponding probability density function

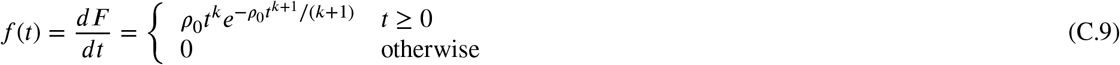

Thus, by computing 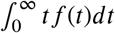, the mean time 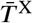 each individual spends in class X is in terms of the Gamma function Γ (⋅) (which is a generalization of the factorial function to real numbers)

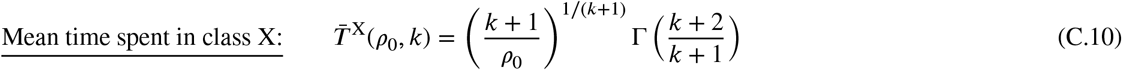

Thus it follows that

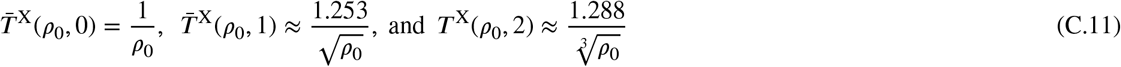

and that 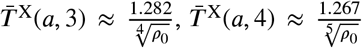, and 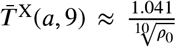. More generally, as a mathematical curiosity, it appears from numerical experiments that 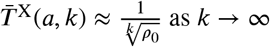.

